# Subgrouping School-Aged Children on the Autism Spectrum Based on Co-Occurring Psychiatric Symptoms

**DOI:** 10.1101/2021.07.19.21260784

**Authors:** Antonia M. H. Piergies, Tomoya Hirota, Rei Monden, Shuting Zheng

## Abstract

**Background:** Phenotypic heterogeneity along the autism spectrum is compounded by co-occurring psychiatric conditions. Deriving subgroups of autistic individuals based on symptoms of these conditions could better our understanding of psychiatric symptom patterns existing within this population. This study’s goals were to derive subgroups of school-aged autistic children based on co-occurring psychiatric symptoms while controlling for age and sex and to examine correlates of subgroup membership while controlling for the degree of ASD-related diagnostic features.

**Method:** Latent class models were estimated in a sample from the Simons Simplex Collection (*n* = 2,087) using “borderline/clinical” versus “normative” range data from five of the *DSM*-Oriented Scales from the CBCL/6-18 as indicator variables. We evaluated the predictive value of NVIQ < 70, atypical sleep duration, allergies/autoimmune conditions, gastrointestinal conditions, and neurological conditions on subgroup membership using multinomial logistic regression.

**Results:** Four subgroups emerged: *Low Psychiatric Symptoms* (41.02%), *Externalizing Symptoms* (12.36%), *Internalizing Symptoms* (31.58%), and *High Psychiatric Symptoms* (15.05%). Key findings were that NVIQ < 70 was associated with decreased odds of belonging to the *Internalizing Symptoms* and *High Psychiatric Symptoms* subgroups over the *Low Psychiatric Symptoms* subgroup, while atypical sleep duration and gastrointestinal conditions were associated with increased odds of belonging to the *Externalizing Symptoms* and *High Psychiatric Symptoms* subgroups. Neurological conditions were also associated with increased odds of belonging to the *Externalizing Symptoms* subgroup.

**Conclusion:** Distinct patterns of psychiatric symptoms exist within school-aged autistic children and are correlated with NVIQ < 70, atypical sleep duration, and medical conditions, providing insights for clinical practice and etiology-driven research.

## Introduction

Co-occurring psychiatric conditions are prevalent among autistic children (Rosen et al., 2018), with 70-91% meeting diagnostic criteria for at least one at some point during their lifetimes (Leyfer et al., 2006; Simonoff et al., 2008; Salazar et al., 2015; Mosner et al., 2019). Common co-occurring conditions include anxiety disorders, attention-deficit/hyperactivity disorder (ADHD), depressive disorders, obsessive-compulsive disorder (OCD), and oppositional defiant disorder (ODD; Simonoff et al., 2008; Gjevik et al., 2011; Lai et al., 2019).

The presence of psychiatric conditions has been linked to increased impairments in adaptive, social, and academic functioning (Kim et al., 2000; Sikora et al., 2012; Kaat et al., 2013), and can make diagnosing autism spectrum disorder (ASD) more challenging (Levy et al., 2010; Rosen et al., 2018). For example, the presence of internalizing and externalizing symptoms—those that are internally- (e.g., anxiety problems) and externally-driven (e.g., conduct problems), respectively—has been shown to impact the sensitivity and specificity of commonly-used ASD screeners and diagnostic measures, including the Autism Diagnostic Interview-Revised (ADI-R), the Autism Diagnostic Observation Schedule (ADOS), the Modified Checklist for Autism in Toddlers (MCHAT), and the Social Responsiveness Scale (SRS; Havdahl et al., 2016; Colombi et al., 2019; Christopher et al., 2020). Additionally, the presence of internalizing and externalizing symptoms appear to differentially affect intervention responses in autistic children. For example, one study reported that autistic children with ADHD did not respond to a ten-week social skills training, whereas those with an anxiety disorder and those without a co-occurring psychiatric condition did (Antshel et al., 2011).

Co-occurring psychiatric conditions further contribute to the phenotypic heterogeneity of ASD. On the one hand, they can influence the presentation of core ASD diagnostic features (Wood & Gadow, 2010; Sprenger et al., 2013) and the presentation of associated psychiatric symptoms (Sinzig et al., 2008). For example, compared to autistic children without ADHD and children with ADHD alone, autistic children with ADHD can present with more attentional and social difficulties (Sinzig et al., 2008; Sprenger et al., 2013), and autistic children with an anxiety or a depressive disorder can present with more restricted/repetitive behaviors and social withdrawal (Chandrasekhar & Sikich, 2015; Muskett et al., 2019). On the other hand, co-occurring psychiatric conditions can make associated symptoms, such as anxiety, present atypically (Kerns & Kendall, 2012). In autistic children, anxiety can manifest as obsessive-compulsive behaviors without premonitory distress and uncommon types of specific phobia, for example. Since some psychiatric conditions are more likely to present together in the same, autistic child than others—for example, an ADHD diagnosis is often accompanied by diagnoses of an anxiety disorder, a mood disorder, or ODD (Gadow et al., 2008a; Gordon-Lipkin et al., 2018)—patterns of psychiatric symptoms are suspected to exist in this population. Deriving subgroups based on co-occurring psychiatric symptoms could facilitate the identification of symptom patterns in this population, which could in turn be used in future studies to better our understanding of the shared and distinct mechanisms underlying specific symptom profiles. Transdiagnostic constructs, in particular, could prove useful to prevention, screening, detection, and intervention efforts and lead to more efficient delivery of services. For example, shifting the focus of psychiatric symptom measures away from diagnostic categories and towards constructs that are impaired across disorders would allow for interventions that are both more tailored to one’s needs and more generalizable along the autism spectrum, as autistic children often present with multiple, co-occurring conditions (Simonoff et al., 2008).

One useful technique for deriving subgroups is latent class analysis (LCA; Porcu & Giambona, 2017), a person-centered approach that assigns individuals to otherwise unobserved classes based on a combination of observed categorical variables. Several studies have used person-centered approaches when attempting to disentangle the heterogeneity in the expression of autistic traits (Gotham et al., 2012; Lord et al., 2012; James et al., 2016; Lerner et al., 2017; Kim et al., 2019; Harris et al., 2021) and the presentation of other developmental and behavioral characteristics (Landa et al., 2012; Wiggins et al., 2017; Kang et al., 2020; Zheng et al., 2020). However, relatively little attention in the subgrouping literature has been given to co-occurring psychiatric symptoms, which have been recognized as an area of importance (Lombardo et al., 2019). The few studies that have considered co-occurring psychiatric symptoms when subgrouping samples of autistic children are limited in that they either focused on older, school-aged children (≥ 15 years) and aggregated all psychiatric symptoms into a single large category (Doshi-Velez et al., 2014) or focused on younger, preschool-aged children and kept psychiatric symptoms separated, even though multiple, co-occurring conditions are more common in older children (Wiggins et al., 2017; Dovgan & Mazurek, 2019; Nordahl et al., 2020). Additionally, all these studies included other child-level indicator variables (e.g., adaptive functioning, cognitive development) in their models along with psychiatric symptoms. Although these studies have demonstrated the potential for identifying more homogeneous ASD subgroups, no study has systematically examined whether meaningful subgroups of children within the entire school-aged range (6 to 18 years) can be formed based solely on patterns of psychiatric symptoms. The school-aged period is crucial to the study of psychiatric symptoms, as most psychiatric conditions emerge within it as children undergo physiological and neurodevelopmental changes (e.g., puberty) and are exposed to a myriad of environmental stimuli (e.g., interactions with caregivers, peers, and teachers; Solmi et al., 2021).

The presentation of co-occurring psychiatric conditions in autistic children is complex and can be influenced by various factors, such as cognitive ability, sleep duration, and medical history. Regarding cognitive ability, some studies show that autistic children with below average cognitive ability experience more anxiety and exhibit more challenging behaviors (e.g., aggression, self-injury; Bradley et al., 2004; O’Brien & Pearson, 2004; Murphy et al., 2009), while others suggest more internalizing and externalizing symptoms in those with average or above average cognitive ability (Gadow et al., 2005; Gadow et al., 2008b; van Steensel et al. 2011; Gjevik et al., 2011; Strang et al., 2012). With regard to sleep duration, atypical sleep duration, especially that due to insomnia, has been associated with increased internalizing and externalizing symptoms, namely attention difficulties, anxiety, and mood dysregulation, in autistic children (Park et al., 2012; Richdale & Baglin, 2015; Veatch et al., 2017).

In addition to co-occurring psychiatric symptoms, it is estimated that 10-77% of autistic children have a co-occurring medical condition, with gastrointestinal conditions, neurological conditions, and allergies/autoimmune conditions being among the most well-studied (Muskens, et al., 2017). The presence of medical conditions, especially gastrointestinal and neurological conditions, has also been positively associated with internalizing and externalizing symptoms in autistic children (Gadow et al., 2008b; Simonoff et al., 2008; Mannion & Leader, 2013; Mazefsky et al., 2014; Fulceri et al., 2016; Weber & Gadow, 2017; Restrepo et al., 2020). However, several studies have not found significant associations between other medical conditions (e.g., allergies/autoimmune conditions) and co-occurring psychiatric symptoms in autistic children (Ming et al., 2008; Weber & Gadow, 2017). Considering these mixed findings, more research is needed to investigate the relationship between child-level factors and co-occurring psychiatric symptoms, especially in terms of symptom-based subgroups.

In addition, age and sex have been shown to differentially affect the presentation of co-occurring psychiatric symptoms in autistic children. Older children seem to be at higher risk for internalizing and externalizing symptoms than younger children (Fodstad et al., 2010; Salazar et al., 2015; Soke et al., 2018), while internalizing symptoms are more often reported in females, and externalizing symptoms are more often reported in males (Giarelli et al. 2010; Solomon et al., 2012; Salazar et al., 2015). These studies imply that age and sex should be carefully controlled during investigations of co-occurring psychiatric symptoms in autistic children. Lastly, there is emerging but inconsistent evidence of a relationship between the degree of ASD-related diagnostic features and co-occurring psychiatric symptoms, cognitive ability, atypical sleep duration, and medical conditions, with studies reporting positive (Yoshida & Uchiyama, 2004; Jang et al., 2010; Snow & Lecavalier, 2011; Wang et al., 2011; Ewen et al., 2019; Lindor et al., 2019) and negative (Mosner et al., 2020) relationships as well as no relationship (Simonoff et al., 2013; Louwerse et al., 2015; Lindor et al., 2019). Therefore, studies probing whether these factors influence the presentation of co-occurring psychiatric symptoms should control for the degree of ASD-related diagnostic features.

The purpose of this study was (1) to derive subgroups of school-aged autistic children based on the presence of co-occurring psychiatric symptoms while controlling for age and sex and (2) to examine predictors of subgroup membership, focusing on the presence of cognitive ability, atypical sleep duration, and three groups of medical conditions—allergies/autoimmune conditions, gastrointestinal conditions, and neurological conditions—while controlling for the degree of ASD-related diagnostic features.

## Methods

### Participants

Data were obtained from the Simons Simplex Collection (SSC; version 15). The data collection process has been described in detail elsewhere (Fischbach & Lord, 2010) and briefly summarized below. Phenotypic data were collected over a six-month period from more than 2,700 families that were already receiving services from one of twelve university-affiliated clinics. Each family was composed of a single autistic child (i.e., the proband), their unaffected parents, and their unaffected sibling(s). To be included in the SSC, probands needed to be English-speaking, older than 4 years old, and younger than 18 years old. Probands also needed to have a best-estimate diagnosis of ASD and meet designated criteria on the ADI-R, the ADOS, and a measure of nonverbal cognitive ability. Specifically, for the ADI-R, probands were required to either surpass standard cutoffs on both the Language/Communication and Reciprocal Social Interactions domains, surpass one domain’s cutoff and be within two points of the other domain’s cutoff, or be within one point of both domains’ cutoffs. For the ADOS, probands were required to surpass cutoffs for Autism Spectrum or Autism classification. For the measure of nonverbal cognitive ability, probands under 6 years and 11 months of age were required to have a nonverbal mental age ≥ 24 months, although exceptions of mental ages < 24 months but ≥ 18 months were made after careful review when necessary. Probands 7 years of age or older were required to have a nonverbal mental age of ≥ 30 months (see the SSC’s Researcher Welcome Packet for more information on inclusion criteria). Informed consent was obtained at each data collection site included in the SSC.

Figure 1 illustrates the participant selection process for the current study. Our original analytic sample consisted of 2,184 school-aged autistic children (aged 6 to 18 years old) whose caregivers had completed a CBCL/6-18. For the LCA, 97 (4.44%) of the participants were excluded because they had missing data for indicator variables and/or covariates (described below), resulting in a sample of 2,087 (95.56%) children. Further, 36 (1.72% of 2,087) participants from the LCA sample were excluded from the multinomial logistic regression due to missing indicator variables data, resulting in a sample of 2,051 (93.91%) children. An additional 97 (4.73% of 2,051) participants were excluded from this analysis because they were considered outliers, having standardized residuals > |2| and therefore violating the assumptions of multinomial logistic regression. Thus, the final sample consisted of 1,954 (89.47%) children.

**Figure 1.**
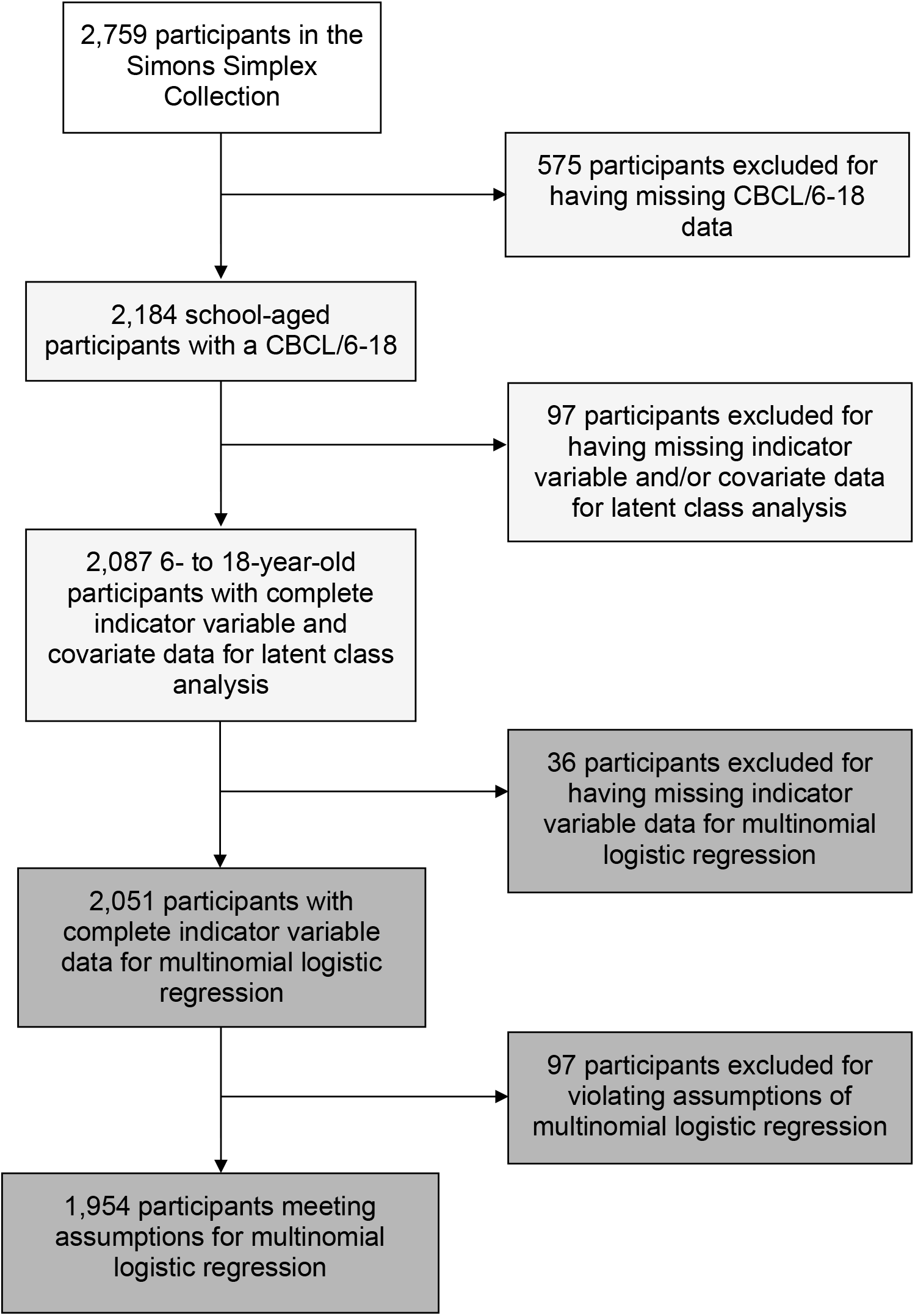
Flowchart illustrating the participant selection process for all analyses.

### Measures

#### Indicator Variables for Latent Class Analysis

The presence of co-occurring psychiatric symptoms was determined using the *DSM*-Oriented Scales of the caregiver-report form of the CBCL/6-18. The scales are comprised of items that describe specific emotions and behaviors. These items are first rated 0 (“not true”) to 2 (“often” or “very true”) by caregivers. *T*-scores are then calculated based on raw score totals. While the *DSM*-Oriented Scales are more dimensional measures of symptom levels (although *T*-scores may not even capture the full range of variation), for the purpose of the current analysis, we dichotomized *T*-scores based on the recommended cutoffs separating the “borderline/clinical” (elevated levels of symptoms defined by *T*-scores ≥ 65) and “normative” ranges (absence of elevated symptoms defined by *T*-scores < 65; Achenbach & Rescorla, 2001). We estimated latent class models using five of the six *DSM*-Oriented Scales: ADHD Problems, Affective Problems, Anxiety Problems, Conduct Problems, and Oppositional Defiant Problems. We excluded the Somatic Problems scale from our analysis given previously-reported concerns around its psychometric properties (Ebesutani et al., 2009) and its overlap with other scales. We also conducted a sensitivity analysis, using all six scales as indicator variables in our LCA and found little added value when the Somatic Problems scale was included (Supplementary Table 1; Supplementary Figure 1) and observed substantial agreement between class assignments from LCAs with and without the Somatic Problems scale (kappa > 0.80; Cohen, 1960).

#### Indicator Variables for Multinomial Logistic Regression

##### Nonverbal IQ

Nonverbal IQ (NVIQ) was assessed using one of four measures included in the SSC data collection process, the Mullen Scales of Early Learning (Mullen, 1995), the Differential Ability Scales-II (Elliott, 2007), the Wechsler Intelligence Scale for Children-IV, or the Wechsler Abbreviated Scale of Intelligence (Wechsler, 1999; Weschler, 2003). Of note, for the Mullen, NVIQ was calculated by averaging age-equivalent scores from the Fine Motor and Visual Reception scales, dividing this average by the child’s chronological age, and multiplying this quotient by 100 (Bishop et al., 2011).

##### Atypical sleep duration

The classification of sleep duration as atypical was determined by following the methods of a previous SSC study (Veatch et al., 2017); children whose current sleep duration fell outside of the recommended age-based ranges, per the American Academy of Pediatrics (Paruthi et al., 2016), were considered to have sleep durations that were atypical. A manually-cleaned version of this variable has been made available (Supplementary Table 8).

##### Medical conditions

Information about medical history was obtained from the SSC’s medical history form, indicating whether or not participants had ever had a given condition, as reported by their caregiver(s). We grouped conditions into larger categories based on the organization of the SSC’s medical history form and a previous study (Croen et al., 2015; Supplementary Tables 2 and 3). While we started with nine groups in total—*Allergies/Autoimmune Conditions, Cancer, Chronic Illnesses, Diseases & Surgeries, Gastrointestinal Conditions, Genetic Conditions, Hearing and Vision Problems, Heart Disease*, and *Neurological Conditions*—we ultimately included the three that were most relevant to co-occurring psychiatric symptoms in autistic children based on previous studies: *Allergies/Autoimmune Conditions, Gastrointestinal Conditions*, and *Neurological Conditions* (Muskens et al., 2017; Tye et al., 2019; see Supplementary Table 2 for each group’s endorsement rate). The variables representing these three groups were coded dichotomously, with 1 indicating that a participant had a history of at least one of the conditions within the category (i.e., responses of “true” or “diagnosed” on the SSC’s medical history form) and 0 indicating that a participant did not have a history of any condition within the category (i.e., responses of “false” or “no” on the SSC’s medical history form). Ambiguous responses, such as “not sure” or “suspected,” were also coded as 0. If all the conditions in the category were missing responses, the category was coded as “NA.” The conditions in all three groups are displayed in Supplementary Table 3. The individual conditions with the highest endorsement rates in each category were: allergies and/or reactions to the environment (27.31%), food (14.37%), and medication (13.90%), asthma (10.30%), constipation (18.30%), diarrhea (6.71%), gastroesophageal reflux (6.61%), unusual stools (5.56%), excessively clumsy/uncoordinated due to a neurological condition (8.10%), and seizures (5.61%).

#### Covariates

Age was approximated using participants’ age at ADOS administration. The degree of ASD-related diagnostic features was captured using the ADOS’s Comparison Scores (CSS; Gotham et al., 2009). Because CSS data could not be calculated for Module 4 participants at the time of collection, and were thus missing, we calculated CSS data for Module 4 participants based on the revised algorithm and added these data to our analysis (Hus & Lord, 2014).

### Statistical Analyses

LCA was performed in R (version 4.0.4) using the poLCA package (Linzer & Lewis, 2013). Latent class models with one through six classes were estimated based on five binary indicator variables (from the CBCL/6-18’s *DSM*-Oriented Scales): ADHD symptoms, affective symptoms, anxiety symptoms, conduct symptoms, and oppositional defiant symptoms. Age and sex were included in the models as covariates. Comparisons between models were made using goodness-of-fit criteria, including the Akaike information criterion (AIC), the Bayesian information criterion (BIC), and entropy. Lower AIC and BIC values and higher entropy values indicate better model fit. Additional goodness-of-fit criteria (e.g., Gsq, Llik, cAIC, aBIC) are reported in Supplementary Table 4. Models were also evaluated for interpretability (i.e., whether the derived classes were clinically relevant and distinguishable). Posterior probabilities were used to assign participants to their most likely class. Multinomial logistic regression was performed in SPSS (version 27; IBM Corporation, Armonk, NY) to investigate whether the derived classes could be predicted by five variables—nonverbal IQ (NVIQ) < 70, atypical sleep duration, allergies/autoimmune conditions, gastrointestinal conditions, or neurological conditions—while controlling for the degree of ASD-related diagnostic features (measured by ADOS CSS data). Residual analysis was used to check relevant assumptions: linearity, normality, and homogeneity of variance. Multicollinearity was checked using Cramer’s V. All analysis scripts can be found here: https://osf.io/xyvc2/?view_only=736ea37d02404383bc9125745e3c2391.

## Results

For the entire LCA sample (*n* = 2,087), the mean age was 10.30 years (range: 6-18, *SD* = 3.12) and 1,808 (86.63%) participants were male (Table 1). Additionally, the sample included 516 (24.72%) participants who had NVIQs < 70 (*M* = 84.48, *SD* = 26.28), 501 (24.01%) participants who had atypical sleep duration, 992 (47.53%) who had histories of at least one allergy/autoimmune condition, 664 (31.82%) who had histories of at least one gastrointestinal condition, and 517 (24.77%) who had histories of at least one neurological condition.

**Table 1.** Demographic and clinical characteristics of all four latent classes.

| Sample Descriptives | Low Psychiatric Symptoms |  | Externalizing Symptoms |  | Internalizing Symptoms |  | High Psychiatric Symptoms |  | Total Sample |  |
| --- | --- | --- | --- | --- | --- | --- | --- | --- | --- | --- |
|  | n | % | n | % | n | % | n | % | n | % |
| Sample Size | 856 | 41.02 | 258 | 12.36 | 659 | 31.58 | 314 | 15.05 | 2,087 | 100 |
| Sex (males) | 723 | 84.46 | 195 | 75.58 | 611 | 92.72 | 279 | 88.85 | 1,808 | 86.63 |
| Age<sup>a</sup> (years) | 10.14 | 3.23 | 9.47 | 2.73 | 10.93 | 3.14 | 10.11 | 2.85 | 10.30 | 3.12 |
| ADOS CSS<sup>a</sup> | 7.49 | 1.72 | 7.38 | 1.69 | 7.52 | 1.70 | 7.36 | 1.66 | 7.47 | 1.70 |
| NVIQ<sup>a</sup> | 82.88 | 26.76 | 84.53 | 27.83 | 87.08 | 26.17 | 85.02 | 25.62 | 84.73 | 26.59 |
| Indicator Variables |  |  |  |  |  |  |  |  |  |  |
| ADHD Problems | 138 | 16.12 | 146 | 56.59 | 267 | 40.52 | 256 | 81.53 | 807 | 38.67 |
| Affective Problems | 52 | 6.07 | 108 | 41.86 | 369 | 56.00 | 280 | 89.17 | 809 | 38.76 |
| Anxiety Problems | 44 | 5.14 | 22 | 8.53 | 559 | 84.83 | 311 | 99.04 | 936 | 44.85 |
| Conduct Problems | 0 | 0 | 162 | 62.79 | 3 | 0.46 | 241 | 76.75 | 406 | 19.45 |
| Oppositional Defiant Problems | 0 | 0 | 177 | 68.60 | 88 | 13.35 | 279 | 88.85 | 544 | 26.07 |
| NVIQ < 70 | 239 | 27.92 | 68 | 26.36 | 141 | 21.40 | 68 | 21.66 | 516 | 24.72 |
| Atypical Sleep Duration | 178 | 20.79 | 64 | 24.81 | 166 | 25.19 | 93 | 29.62 | 501 | 24.01 |
| Allergies/Autoimmune Conditions | 376 | 43.93 | 121 | 46.90 | 332 | 50.38 | 163 | 51.91 | 992 | 47.53 |
| Gastrointestinal Conditions | 232 | 27.10 | 89 | 34.50 | 227 | 34.45 | 116 | 36.94 | 664 | 31.82 |
| Neurological Conditions | 183 | 21.38 | 72 | 27.91 | 176 | 26.71 | 86 | 27.39 | 517 | 24.77 |

<sup>a</sup> Age, ADOS CSS, and NVIQ are described using mean and standard deviation rather than number (n) and percent (%).

### Latent Class Analysis

The results of the LCA (Table 2), which controlled for age and sex, showed that a four-class model was optimal, with the smallest AIC and BIC. None of the entropy values were close to zero, indicating that classes were well-separated (Zhang et al., 2018). The four-class model (Figure 2) showed clear differentiation between co-occurring psychiatric symptom patterns among autistic children and adolescents, the characteristics of which are summarized in Table 3. We also performed an LCA which repeated model estimation 1,000 times using a subset of 1,000 participants as a validation sample, which suggested either a three- or four-class solution (Supplementary Table 4, Figure 2). The *Low Psychiatric Symptoms* subgroup was characterized by the lowest percentage of participants with “borderline/clinical” levels of all symptoms. Compared to the *Low Psychiatric Symptoms* subgroup, the *Externalizing Symptoms* subgroup had more participants with “borderline/clinical” levels of all symptoms, mainly ADHD, conduct, and oppositional defiant symptoms. The *Internalizing Symptoms* subgroup had fewer participants with “borderline/clinical” levels of ADHD, conduct, and oppositional defiant symptoms but more participants with “borderline/clinical” levels of affective and anxiety symptoms than the *Externalizing Symptoms* subgroup. The *High Psychiatric Symptoms* subgroup was characterized by the highest percentage of participants with “borderline/clinical” levels of all symptoms.

**Table 2.** Goodness-of-fit criteria for all six latent class models.

| Model | *df* | AIC | BIC | Entropy | Smallest Class (%) |
| --- | --- | --- | --- | --- | --- |
| 1-class | 26 | 12,904.46 | 12,932.68 | 3.09 | NA |
| 2-class | 18 | 11,609.27 | 11,682.64 | 2.80 | 34.61 |
| 3-class | 10 | 11,495.77 | 11,614.28 | 2.75 | 25.97 |
| 4-class | 2 | 11,440.13 | 11,603.79 | 2.74 | 14.55 |
| 5-class | -6 | 11,473.61 | 11,682.42 | 2.74 | 0 |
| 6-class | -14 | 11,611.02 | 11,864.98 | 2.77 | 0 |

*Note.* Latent class analysis was conducted with five *DSM*-Oriented Scales of the Child Behavior Checklist for Ages 6 to 18 (Anxiety Problems, Affective Problems, Attention Deficit/Hyperactivity Problems, Conduct Problems, and Oppositional Defiant Problems) as indicator variables. Age and sex were used as covariates.

**Table 3.** Odds ratios (OR) and 95% confidence intervals (95% CI) describing the relationship between indicator variables and latent class membership.

| Indicator Variables | Externalizing Symptoms<br>(n = 189) |  | Internalizing Symptoms<br>(n = 648) |  | High Psychiatric Symptoms<br>(n = 274) |  |
| --- | --- | --- | --- | --- | --- | --- |
|  | OR | 95% CI | OR | 95% CI | OR | 95% CI |
| Allergies/Autoimmune Conditions | 1.43* | 1.03-1.98 | 1.27* | 1.03-1.57 | 1.46** | 1.10-1.93 |
| Atypical Sleep Duration | **2.07\*\*\*** | 1.45-2.94 | 1.37* | 1.07-1.75 | **2.15\*\*\*** | 1.58 -2.93 |
| Gastrointestinal Conditions | **2.11\*\*\*** | 1.51-2.94 | 1.39** | 1.11-1.75 | **1.90\*\*\*** | 1.41-2.55 |
| Neurological Conditions | **2.15\*\*\*** | 1.52-3.05 | 1.39** | 1.09-1.77 | 1.58** | 1.15-2.17 |
| NVIQ < 70 | .72 | .49-1.05 | **.63\*\*\*** | .49-.81 | **.42\*\*\*** | .29-.61 |

*Note.* The *Low Psychiatric Symptoms* subgroup was compared to the *Externalizing Symptoms*, *Internalizing Symptoms*, and *High Psychiatric Symptoms* subgroups. For each indicator variable, the reference group was 0, meaning that participants did not have a NVIQ < 70, atypical sleep duration, or a history of at least one gastrointestinal, neurological, or allergy/autoimmune condition. The degree of ASD-related diagnostic features, measured by ADOS CSS data, was used as a covariate. 97 outliers were excluded for violating the assumptions of multinomial logistic regression.
\*p<.05. \*\*p<.01. \*\*\*p <.001.

**Figure 2.**
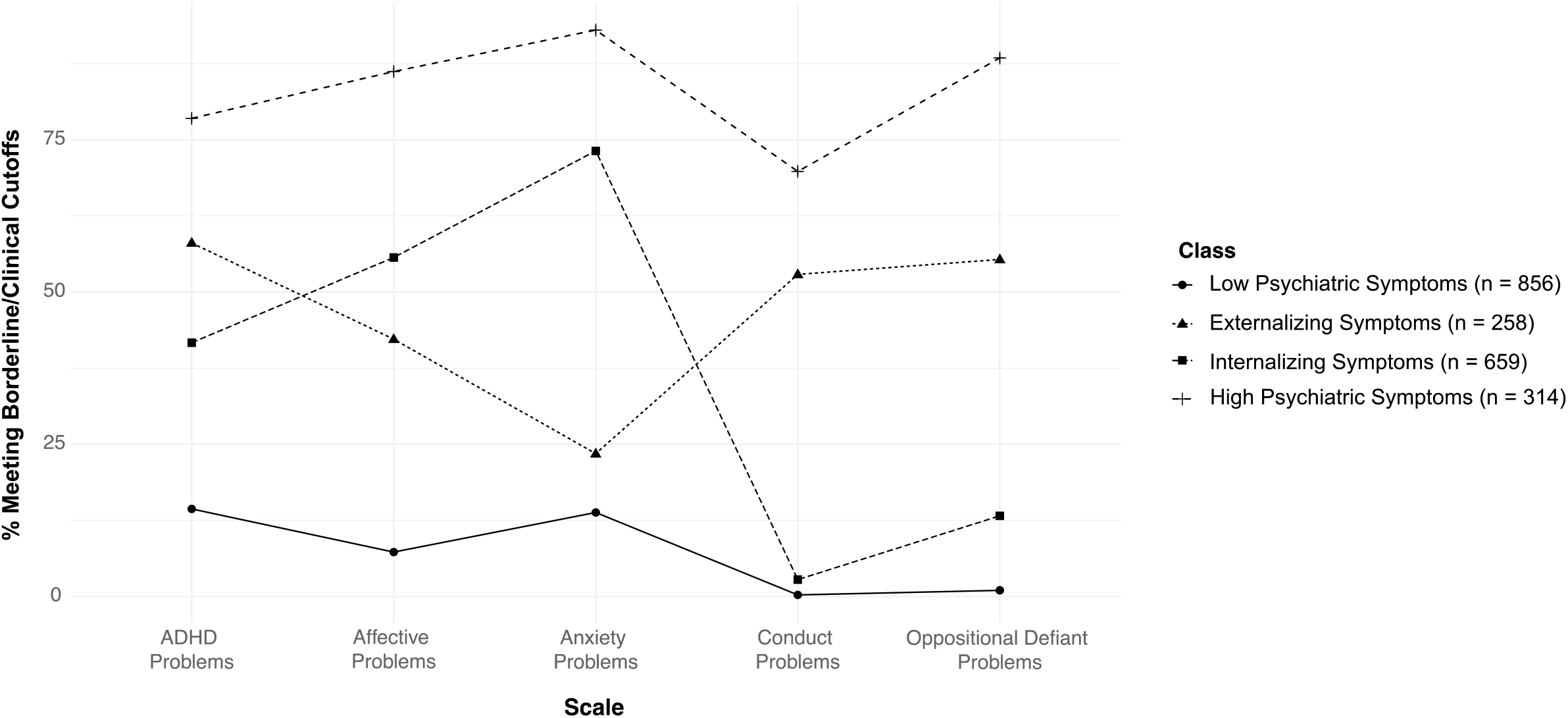
Percent of participants in each latent class with borderline/clinical levels of co-occurring psychiatric symptoms. *Note*. Latent class analysis was conducted with five *DSM*-Oriented Scales of the Child Behavior Checklist for Ages 6 to 18 (Anxiety Problems, Affective Problems, Attention-Deficit/Hyperactivity Problems, Conduct Problems, and Oppositional Defiant Problems) as indicator variables. Age and sex were used as covariates.

### Multinomial Logistic Regression

The results of the multinomial logistic regression are presented in Table 3. Comparisons were made while controlling for the degree of ASD-related diagnostic features and using the *Low Psychiatric Symptoms* subgroup as the reference. NVIQ < 70 (*p* < .001), atypical sleep duration (*p* < .001), allergies/autoimmune conditions (*p* < .05), gastrointestinal conditions (*p* < .001), and neurological conditions (*p* < .001) were all found to be significant predictors of subgroup membership. Moreover, almost all these variables were significantly associated with belonging to the three subgroups characterized by elevated “borderline/clinical” levels of symptoms. Given discourse surrounding the relationship between statistical significance (i.e., *p*-values) and sample size (Lantz et al., 2012), we chose to focus on predictors with the most robust effects. NVIQ < 70, atypical sleep duration, gastrointestinal conditions, and neurological conditions were the most robust predictors of subgroup membership, making participants approximately two times more or less likely to belong to one of these subgroups. Relative to the *Low Psychiatric Symptoms* subgroup, children with NVIQs < 70 were less likely to belong to the *Internalizing Symptoms* (OR = .63, *p* = < .001) or the *High Psychiatric Symptoms* (OR = .42, *p <* .001) subgroups. Children sleeping shorter or longer than recommended and children with histories of at least one gastrointestinal condition were more likely to belong to the *Externalizing Symptoms* (OR = 2.07, *p* = < .001; OR = 2.11, *p* = < .001) and *High Psychiatric Symptoms* (OR = 2.15, *p* = < .001; OR = 1.90, *p* = < .001) subgroups than the *Low Psychiatric Symptoms* subgroup, whereas children with histories of at least one neurological condition were more likely to belong to the *Externalizing Symptoms* subgroup alone (OR = 2.15, *p* = < .001).

## Discussion and Implications

### Discussion

The current study is the first to use LCA to derive subgroups of school-aged autistic children based on patterns of co-occurring psychiatric symptoms while controlling for age and sex. We identified four subgroups, characterized by low psychiatric symptoms, externalizing symptoms, internalizing symptoms, and high psychiatric symptoms across CBCL scales. Using multinomial logistic regression, while controlling for the degree of ASD-related diagnostic features, we also found evidence of the predictive value of NVIQ < 70, atypical sleep duration, and three groups of medical conditions— allergies/autoimmune conditions, gastrointestinal conditions, and neurological conditions—for subgroup membership.

The *Low Psychiatric Symptoms* subgroup had the lowest endorsement of ADHD, affective, anxiety, conduct, and oppositional defiant symptoms. The *Low Psychiatric Symptoms* subgroup (41.02%) was the largest subgroup in our sample, suggesting that slightly less than half of school-aged autistic children are within the “normative” range for most, if not all, psychiatric symptoms measured by the CBCL/6-18. This rate is similar to rates of “no psychiatric diagnosis” in other ASD samples (37.2%; Simonoff et al., 2008) and is substantially lower than rates of belonging to the equivalent subgroup in general-population and clinically-referred samples, which are presumably majority nonautistic, of past studies that have derived subgroups based on psychiatric symptoms: 62.5% (Olino et al., 2012), 66.32% (Bianchi et al., 2017), 77.41% (Essau & de la Torre-Luque, 2019), 79.2% (Parviainen et al., 2020). Thus, this finding supports the idea of elevated, clinically-relevant psychiatric symptoms in autistic children. Additionally, the most endorsed symptom in our *Low Psychiatric Symptoms* subgroup was ADHD (16.12%), whereas the most endorsed symptoms in the equivalent subgroup of comparison samples were major depressive disorder (40.3%; Olino et al., 2012) and specific phobia (12%; Essau & de la Torre-Luque, 2019). There were negligible differences in endorsement across symptoms in the remaining comparison samples’ equivalent subgroups (Bianchi et al., 2017; Parviainen et al., 2020).

Relative to the *Low Psychiatric Symptoms* subgroup, the *Internalizing Symptoms* subgroup had more symptom endorsement across all five scales, especially for affective and anxiety symptoms. The *Internalizing Symptoms* subgroup was our second largest subgroup (31.58%), which is unsurprising given that anxiety (44.85%) and affective (38.76%) symptoms were the most endorsed psychiatric symptoms in our sample. While the percentage of participants belonging to this subgroup is far greater than the relative size of comparison samples’ equivalent subgroups (9.1-16.9%; Olino et al., 2012; Parviainen et al., 2020), these rates are lower than previously reported rates of anxiety (56%) and depression (44%) in autistic children (Strang et al., 2012). This discrepancy could be due to the fact that 15.05% of our sample belonged to the *High Psychiatric Symptoms* subgroup, which is characterized by high rates of both internalizing and externalizing symptoms. Overall, the presence of a larger subgroup characterized by internalizing symptoms in our sample is in agreement with past studies reporting higher rates of anxiety and depression in autistic individuals as compared to individuals in the general population (White et al., 2009; Kim & Lecavalier, 2021).

The *Externalizing Symptoms* subgroup (12.3%) was the smallest subgroup in our sample, characterized by multiple highly-endorsed externalizing symptoms: ADHD (56.59%), conduct (62.79%), and oppositional defiant (68.60%) symptoms. The *Externalizing Symptoms* subgroup also had more symptom endorsement across all five scales relative to the *Low Psychiatric Symptoms* subgroup but less endorsement of affective and anxiety symptoms and more endorsement of ADHD, conduct, and oppositional defiant symptoms than the *Internalizing Symptoms* subgroup. This subgroup is larger than equivalent subgroups in Essau and de la Torre-Luque’s (2019; 6.97%) and Parvianien and colleagues’ (2020; 9.1%) studies, but smaller than the equivalent subgroup in Olino and colleagues’ study (2012; 16.4%). Participants in equivalent subgroups frequently endorsed multiple symptoms that were indicative of externalizing conditions, including conduct disorder, ODD, and substance use disorders, but not ADHD, although ADHD symptoms were not assessed in one of the three studies. Meanwhile, Bianchi and colleagues’ (2017) study lacks such a subgroup. Rather, their study identified an “ADHD” subgroup, in which ∼70% of children had attention-related symptoms in the “borderline/clinical” range but relatively few other symptoms that were externalizing. Therefore, the externalizing symptoms of autistic children, marked by a combination of ADHD, ODD, and CD, might be more intricate than those in other populations of children.

The *High Psychiatric Symptoms* subgroup had the highest endorsement of all psychiatric symptoms assessed. Notably, this subgroup (15.05%) was about double the size of Bianchi and colleagues’ “Severe Dysregulated” subgroup (7.82%), and substantially larger than the equivalent subgroups in Olino and colleagues’ (2012) and Parviainen and colleagues’ (2020) samples, to which 2.6-4.2% of participants belonged. Essau and de la Torre-Luque’s (2019) sample lacked such a subgroup entirely. Our findings demonstrate an increased rate of complex psychiatric symptom patterns in autistic children as compared to nonautistic children, with symptoms of ADHD often co-occurring with other externalizing and internalizing symptoms. Taken together, the latent structure of co-occurring psychiatric symptoms appears to be similar among autistic and nonautistic children, with the same transdiagnostic factors of “internalizing” and “externalizing” generally explaining symptom co-occurrence in both groups (Rodriguez-Seijas et al., 2020). Our study also appears to support the idea that symptom clusters (e.g., those involving ADHD symptoms) may result from a process that is unique to ASD. We observed medium-to-high endorsement rates of ADHD symptoms in the *Externalizing Symptoms, Internalizing Symptoms*, and *High Psychiatric Symptoms* subgroups (range: 40.52-81.53%), in addition to an endorsement rate of 16.12% in the *Low Psychiatric Symptoms* subgroup. This observation is consistent with the literature, as ASD and ADHD co-occur at rates greater than chance (Miller et al., 2019). Having ADHD symptoms might increase autistic children’s risk of experiencing and exhibiting other psychiatric symptoms, such as anxiety, aggression, delinquent behavior, and thought problems (Gadow et al., 2009; Matsushima et al., 2008). Indeed, the most common combination of co-occurring psychiatric conditions in autistic children is ADHD, anxiety disorders, and ODD (Brookman-Frazee et al., 2018).

Multiple, co-occurring psychiatric conditions impact autistic children’s participation in daily activities (Dovgan & Mazurek, 2019). Our subgroups represent the need for interventions targeting aspects of internalizing and externalizing symptoms singly as well as simultaneously. While interventions undoubtedly need to be individualized to align with the priorities of autistic children and their families, identifying a set of educational strategies that are effective for relatively homogeneous subgroups could be feasible, although more research is needed. For example, studies assessing the service needs of autistic children belonging to the *High Psychiatric Symptoms* subgroup could be of great importance, as internalizing symptoms may be overshadowed by externalizing symptoms. Internalizing symptoms affect school performance and peer relationships (Reaven, 2009), yet they might be less obvious targets for parents and teachers and, therefore, may be left unaddressed. One study did not find a relationship between internalizing symptoms and receipt of school services (Rosen et al., 2019). However, externalizing symptoms have been negatively associated with receipt of school services (Rosen et al., 2019). It is speculated that this relationship was due to a higher priority assigned to disruptive behaviors and thus the pursuit of out-of-school interventions. In line with this thought, another study found that autistic children with ADHD were likely to receive out-of-school interventions in addition to in-school interventions (Zablotsky et al., 2020). Thus, the need for research on the adaptation of screeners and evidence-based interventions for the internalizing and externalizing symptoms of autistic children should continue to be emphasized to ensure that sufficient support is available to this population (Kerns et al., 2016; Kerns et al., 2017).

Using multinomial logistic regression, we found that cognitive ability and atypical sleep duration were associated with subgroup membership. Specifically, we found that children with NVIQs < 70 were about half as likely to belong to the *Internalizing Symptoms* or *High Psychiatric Symptoms* subgroups than the *Low Psychiatric Symptoms* subgroup, which is consistent with previous studies suggesting that autistic individuals with average or above average cognitive ability are more often reported to have certain psychiatric conditions, such as anxiety and depressive disorders, than those with below average cognitive ability (Lecavalier, 2006; Hallett et al., 2013a; Sukhodolsky et al. 2008; Mazurek & Kanne 2010). In fact, Mayes and colleagues (2022) recently found that caregivers of autistic children with NVIQs < 70 generally reported lower internalizing and externalizing symptoms in their children than caregivers of autistic children with NVIQs ≥ 70. Specifically, these children had lower oppositional, irritable, tantrum, mean/bullying, lying/cheating, generalized anxiety, and depression ratings but higher separation anxiety ratings. Of note, these results were similar in a sample of children with ADHD (but without ASD). Because NVIQ < 70 was a robust predictor of *Internalizing Symptoms* and *High Psychiatric Symptoms* subgroup membership in our study, our results could be driven by data from participants with NVIQs ≥ 70, who make up over 75% of our sample. Indeed, mean NVIQ was highest for the *Internalizing Symptoms* subgroup (87.08), followed by the *High Psychiatric Symptoms* subgroup (85.02), and then the *Externalizing Symptoms* (84.53) and *Low Psychiatric Symptoms* (82.88) subgroups. It is also possible that difficulties expressing subjective emotional experiences is contributing to an underestimation of caregiver-reported psychiatric symptoms in autistic children with NVIQs < 70 (Simonoff et al., 2008; van Steensel et al., 2017), as the directionality of the relationship between cognitive ability and psychiatric conditions appears to be informant-dependent (Gadow et al., 2008b; Hallet et al., 2013b). Thus, future studies should use alternative strategies to measure psychiatric symptoms in samples of autistic children who vary in cognitive ability. For example, objective measures, such as physiological data, could help clinicians and researchers capture emotional and behavioral responses in autistic children with lower NVIQs (Kushki et al., 2013). Alternatively, autistic children with average or above average cognitive ability may be more aware of accessibility-related challenges, and this awareness could be a risk factor for psychiatric conditions (Sterling et al., 2008). Finally, parents may be overestimating psychiatric symptoms in autistic children with average or above average cognitive ability (Nicpon et al., 2010).

Atypical sleep duration predicted membership to the *Externalizing Symptoms* and *High Psychiatric Symptoms* subgroups. This finding is in line with the results of Adams and colleagues’ (2014) study involving autistic children, in which increased severity of sleep disturbance was positively associated with externalizing and total (i.e., internalizing and externalizing) symptoms, as well as the results of other studies (e.g., Cohen et al., 2018). Insomnia, the more highly endorsed form of atypical sleep duration in our study, has been linked to differences in prefrontal cortex, basal ganglia, and amygdala functioning, which could be impacting executive functioning, reward anticipation, and emotional reactivity in certain contexts, leading to externalizing symptoms or overall high psychiatric symptoms (Maski & Kothare, 2013).

Additionally, using multinomial logistic regression, we found that subgroup membership was predicted by having a history of at least one gastrointestinal condition and a history of at least one neurological condition. This finding complements other studies in the literature that have found a relationship between these medical conditions and any one psychiatric condition in autistic children by adding a novel contribution: demonstrating that a history of these medical conditions predicts unique patterns based on the combined presence/absence of psychiatric symptoms (Gadow et al., 2008b; Simonoff et al., 2008; Mannion & Leader, 2013; Mazefsky et al., 2014; Fulceri et al., 2016; Weber & Gadow, 2017; Restrepo et al., 2020). Our findings are consistent with two studies on how these conditions affect internalizing and externalizing symptoms in autistic children. The first study found more anxiety, externalizing, and total symptoms in young children with gastrointestinal symptoms compared to those without symptoms (Fulceri at al., 2016), which is consistent with our finding that having a history of at least one gastrointestinal condition roughly doubles one’s likelihood of belonging to the *Externalizing Symptoms* and *High Psychiatric Symptoms* subgroups over the *Low Psychiatric Symptoms* subgroup. The second study found that having epilepsy, one of the neurological conditions included in our study, resulted in an increased likelihood of having the psychiatric conditions that correspond to our *Externalizing Symptoms* and *High Psychiatric Symptoms* subgroups (i.e., ADHD, oppositional defiant or conduct disorder, and any emotional disorder) by ∼11- and 17-fold, respectively (Simonoff et al., 2008). In our study, having a history of at least one neurological condition robustly predicted *Externalizing Symptoms* subgroup membership, increasing a child’s likelihood by about two-fold. One possible explanation for the difference in the magnitude of our study’s and this study’s odds ratios is that we included a broader range of neurological conditions (16 conditions, Supplementary Table 3), whereas Simonoff and colleagues focused solely on epilepsy. Possible mediators of this notable association between mental and physical health in autistic children include differences in neural circuitry (e.g., the serotonergic system; Pan et al., 2021), autonomic nervous system functioning (Ferguson et al., 2017), and the gut-brain axis (Chen et al., 2021).

Our study has several limitations. First, we used a categorical framework to conceptualize psychiatric symptoms. Though dichotomizing a dimensional measure (i.e., the *DSM*-Oriented Scales) likely resulted in the loss of information on symptom levels, we chose to do so in order to make our findings directly relevant to clinical practice by focusing on the presence or absence of “borderline/clinical-range” psychiatric symptoms. Second, psychiatric symptoms may present differently in autistic children and/or children with lower cognitive ability (Kerns et al., 2012), and some symptoms may not reflect the same constructs as they do in typically developing (Rodriguez-Seijas et al., 2019) or other clinical populations (Sinzig et al., 2009). Additionally, although the CBCL’s *DSM*-Oriented Scales were designed to map on to a wide range of specific disorders of the *DSM*-5, including major depressive disorder, persistent depressive disorder, generalized anxiety disorder, separation anxiety disorder, social anxiety disorder, specific phobia, ADHD, oppositional defiant disorder, and conduct disorder, they are not all-encompassing. Further, we defined atypical sleep solely based on duration. We also captured the degree of ASD-related diagnostic features via ADOS Comparison Score (CSS) data, which only provide information on behaviors observed in a limited context (Hus et al., 2014). Therefore, our study may have failed to capture some psychiatric symptoms and other co-occurring conditions in this population. Third, the cross-sectional nature of the SSC sample does not allow for the evaluation of causal relationships between our indicator variables and psychiatric symptom-based subgroups. Such relationship is likely bidirectional and warrants longitudinal investigation. Fourth, given the nature of the SSC sample and LCA, our findings may not be widely generalizable to all children on the autism spectrum. With regard to the SSC sample, multiplex families were excluded, inclusion was based on strict ADI-R and ADOS cut-offs, and participants received their best-estimate diagnosis of ASD over a decade ago. Regarding LCA, this method takes a model-based subgrouping approach, and the best fitting model is determined based on the available data, limiting the generalizability to other samples. We also caution fellow clinicians and researchers that our findings should be interpreted with the sampling method in mind, as it may not fully represent the range of psychiatric symptoms in this population due to changing diagnostic criteria and referral processes (Lai et al., 2015; Lord et al., 2018; CDC, 2021). However, our large sample size (∼2,000 autistic individuals) and use of age, sex, and the degree of ASD-related diagnostic features as covariates add to the strengths of our study, which was effective at capturing the individual profiles of co-occurring psychiatric conditions that exist within our sample and predictors of such profiles.

### Implications

The findings of this study could have important implications for clinical practice and research. First, the four subgroups derived from our sample demonstrate a need for clinicians to be cognizant of co-occurring internalizing and externalizing symptoms in school-aged autistic children and the potential impact of these psychiatric symptoms on tools used to detect ASD. In other words, when a clinician is evaluating a school-aged child for ASD, it is likely that the child will be displaying certain patterns of co-occurring psychiatric symptoms that could affect their interpretation of ASD diagnostic measures. Additionally, our subgroups support the idea of initially using broadband screeners to evaluate the mental health of school-aged autistic children before using more targeted, disorder-specific measures, since the majority of participants in our sample had symptoms consistent with more than one condition. Our findings also reinforce the need for multidisciplinary services in this population, suggesting that ASD specialists consider other co-occurring conditions when they are providing care for autistic children. For example, based on our study’s subgroups, when seeing an autistic child with a combination of internalizing and externalizing symptoms, a psychologist may want to collaborate with a physician to monitor for atypical sleep duration, gastrointestinal conditions, and/or neurological conditions. Psychiatric symptom-based subgroups also affect responses to intervention and should be carefully considered in the development of intervention guidelines, which are currently broad (i.e., non-specific to subpopulations), and in future studies on how different subpopulations of autistic children respond to interventions (Antshel et al., 2011; McBride et al., 2020; Dekker et al., 2021; Wickstrom et al., 2021). For example, light therapy could be beneficial to children belonging to a *High Psychiatric Symptoms* subgroup, who are at elevated risk for depression and atypical sleep duration (Fritzsche et al., 2001). Finally, the findings of this study provide additional rationale to investigate the shared and distinct mechanisms between psychiatric conditions and other co-occurring conditions. Relatively little research has been done in this area, but differences in emotion regulation, executive functioning, and cognitive control are among potential candidates (Mazefsky et al., 2013; Neuhaus et al., 2014; Weiss, 2014; Hedley et al., 2021). Since the SSC has extensive phenotypic and genetic data available, future studies could replicate our subgroups and directly examine potential transdiagnostic constructs. For example, several studies have suggested shared genetic contributions to ASD and ADHD phenotypes as well as ASD, depression, and anxiety phenotypes (Ronald et al., 2014; Ruparelia et al., 2017). Studies have also suggested overlap in genetic factors that increase one’s chances of developing ASD, intellectual disability, atypical sleep duration, and medical conditions (Veatch et al., 2015; Jeste & Geschwind, 2014). Finally, studies should investigate contextual factors contributing to the mental health of school-aged autistic children. For example, ASD acceptance from external sources is associated with depression (Cage et al., 2018). In conclusion, our findings indicate that there are unique patterns of psychiatric symptoms among school-aged autistic children which are associated with cognitive ability, atypical sleep duration, and medical conditions. To improve the ways in which we support autistic children, we argue that professionals should go beyond the diagnosis of ASD and consider an individual’s comprehensive profile when diagnosing, treating, and researching this population.

## Data Availability

Approved researchers can obtain the SSC population datasets described in this study by applying at https://base.sfari.org. All the scripts used to run the analyses can be found here: https://osf.io/xyvc2/?view_only=736ea37d02404383bc9125745e3c2391.

https://osf.io/xyvc2/?view_only=736ea37d02404383bc9125745e3c2391

## Acknowledgements

Dr. Monden was partially supported by the Clinical Investigator’s Research Project at Osaka University’s Graduate School of Medicine. Many thanks to Drs. Caroline Lew, Jordan Wickstrom, and Meghan Miller for their valuable feedback on drafts. We are grateful to all of the families at the participating Simons Simplex Collection (SSC) sites, as well as the principal investigators (A. Beaudet, R. Bernier, J. Constantino, E. Cook, E. Fombonne, D. Geschwind, R. Goin-Kochel, E. Hanson, D. Grice, A. Klin, D. Ledbetter, C. Lord, C. Martin, D. Martin, R. Maxim, J. Miles, O. Ousley, K. Pelphrey, B. Peterson, J. Piggot, C. Saulnier, M. State, W. Stone, J. Sutcliffe, C. Walsh, Z. Warren, E. Wijsman). We also appreciate obtaining access to phenotypic data on SFARI Base. Approved researchers can obtain the SSC population datasets described in this study by applying at https://base.sfari.org.

