## Supplementary File for "Subgrouping School-Aged Children on the Autism Spectrum Based on Co-Occurring Psychiatric Symptoms"

This file belongs to the article: "Subgrouping School-Aged Children on the Autism Spectrum Based on Co-Occurring Psychiatric Symptoms" (Piergies et al., *submitted*). It contains results from the four-class-solution latent class analysis which used all six *DSM*-Oriented Scales of the Child Behavior Checklist for Ages 6 to 18 as indicator variables and age and sex as covariates (Supplementary Table 1, Figure 1), frequency tables showing the number and percent of participants with histories of at least one of the medical conditions mentioned in our study (Supplementary Table 2, Table 3), additional goodness-of-fit criteria for the four-class-solution latent class analysis which used five *DSM*-Oriented Scales of the Child Behavior Checklist for Ages 6 to 18 as indicator variables and age and sex as covariates (Supplementary Table 4, Figure 2), results from a sensitivity analysis comparing fit indices between our three- and four-class models which used five *DSM*-Oriented Scales of the Child Behavior Checklist for Ages 6 to 18 as indicator variables and age and sex as covariates after 1,000 repetitions with a subset of 1,000 participants comprising the validation sample (Supplementary Figure 3), a graph illustrating the three-class model which used five *DSM*-Oriented Scales of the Child Behavior Checklist for Ages 6 to 18 as indicator variables and age and sex as covariates (Supplementary Figure 4), frequency tables showing endorsement rates (≥1%) of conditions within medical condition categories and psychiatric symptom-based subgroups (Supplementary Tables 5, 6, and 7), and our manually-cleaned "current sleep duration" variable (Supplementary Table 8).

**Supplementary Table 1**

*Goodness-of-fit criteria for all six latent class models.*

| Model | *df* | AIC | BIC | Entropy | Smallest Class (%) |
|---|---|---|---|---|---|
| 1-class | 57 | 14,676.54 | 14,710.4 | 3.51 | NA |
| 2-class | 48 | 13,252.17 | 13,336.82 | 3.19 | 37.40 |
| 3-class | 39 | 13,103.05 | 13,238.48 | 3.14 | 22.19 |
| 4-class | 30 | 13,039.47 | 13,225.69 | 3.12 | 14.74 |
| 5-class | 21 | 13,049.75 | 13,286.76 | 3.12 | 2.40 |
| 6-class | 12 | 13,223.22 | 13,511.01 | 3.18 | 1.29 |

*Note.* Latent class analysis was conducted with all six *DSM*-Oriented Scales of the Child Behavior Checklist for Ages 6 to 18 (Anxiety Problems, Affective Problems, Attention Deficit/Hyperactivity Problems, Conduct Problems, Oppositional Defiant Problems, and Somatic Problems) as indicator variables. Age and sex were used as covariates.

**Supplementary Figure 1**

*Percent of participants in each latent class with borderline/clinical levels of co-occurring psychiatric symptoms.*

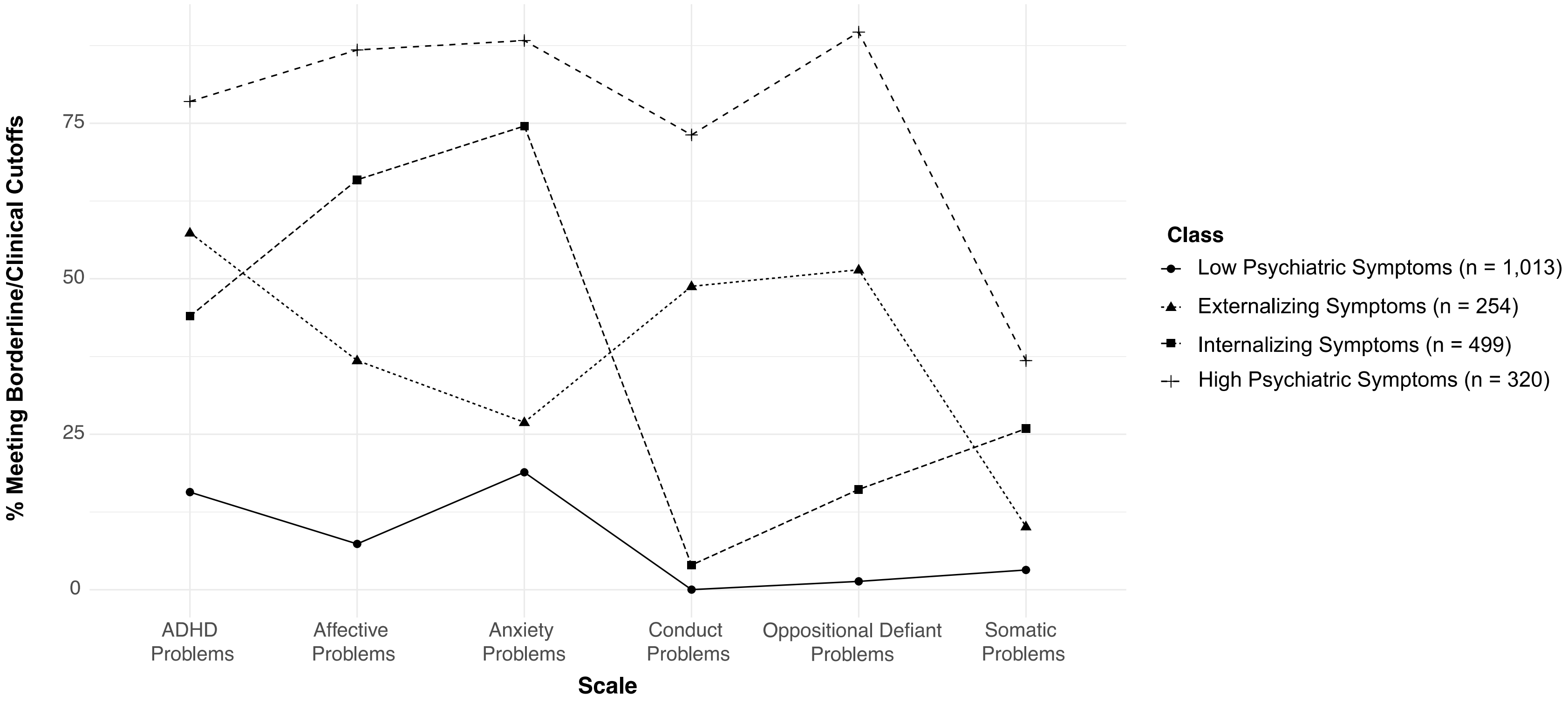

*Note.* Latent class analysis was conducted with all six *DSM*-Oriented Scales of the Child Behavior Checklist for Ages 6 to 18 (Anxiety Problems, Affective Problems, Attention Deficit/Hyperactivity Problems, Conduct Problems, Oppositional Defiant Problems, and Somatic Problems) as indicator variables. Age and sex were used as covariates.

**Supplementary Table 2**

*Number (n) and percent (%) of participants whose caregiver(s) endorsed a history of each medical condition grouped into one of six larger categories: Cancer, Chronic Illnesses, Diseases & Surgeries, Genetic Conditions, Hearing/Vision Problems, and Heart Disease.*

| | Endorsed | |
|---|---|---|
| Broader Category | n | % |
| Cancer | 2 | 0.1 |
| Chronic Illnesses | 30 | 1.44 |
| Diseases & Surgeries | 1,814 | 86.9 |
| Genetic Conditions | 21 | 1 |
| Hearing/Vision Problems | 115 | 5.5 |
| Heart Disease | 4 | 0.19 |

**Supplementary Table 3**

*Number of participants whose caregiver(s) endorsed a history of each medical condition grouped into one of three larger categories: Allergies/Autoimmune Conditions, Gastrointestinal Conditions, and Neurological Conditions.*

| Allergies/Autoimmune Conditions | | | Gastrointestinal Conditions | | | Neurological Conditions | | |
|---|---|---|---|---|---|---|---|---|
| Disease Name | Endorsed | | Disease Name | Endorsed | | Disease Name | Endorsed | |
| | Yes | NA | | Yes | NA | | Yes | NA |
| Adrenal insufficiency (Addison's disease) | 2 | 118 | Bloating (problematic) | 54 | 1,989 | Cerebral palsy | 6 | 2,078 |
| Allergies and/or reactions to medications | 290 | 10 | Constipation (problematic) | 382 | 1,541 | Cranial nerve disorder (e.g., Bell's Palsy) | 2 | 2,085 |
| Asthma | 215 | 143 | Diarrhea (problematic) | 140 | 1,858 | CT/MRI scan done[a] | 40 | 1,576 |
| Bowel disorders | 42 | 122 | Excessive gas (problematic) | 67 | 1,916 | EEG done[a] | 82 | 1,582 |
| Celiac disease | 10 | 117 | Gastric ulcera | 3 | 2,083 | Encephalitis | 2 | 2,085 |
| Diabetes mellitus type I | 1 | 132 | Gastroesophageal reflux | 138 | 1,894 | Excessively clumsy/uncoordinated[b] | 169 | 1,660 |
| Diabetes mellitus type II | 0 | 147 | Inflammatory bowel disease | 2 | 2,081 | Febrile seizures (with fever) | 65 | 2,022 |
| Food allergies | 300 | 8 | Irritable bowel syndrome | 11 | 2,068 | Head injury/loss of consciousness | 86 | 1,963 |
| Environmental allergies | 570 | 9 | Severe abdominal pain | 68 | 1,963 | Meningitis | 8 | 2,076 |
| Hashimoto's thyroiditis | 2 | 120 | Ulcerative colitis | 4 | 2,081 | Movement abnormalities (e.g., tremor) | 34 | 2,014 |
| Hyperthyroidism | 2 | 128 | Unusual stools (problematic) | 116 | 1,874 | PET scan done[a] | 1 | 2,072 |
| Hypothyroidism | 11 | 140 | Vomiting (problematic) | 68 | 1,973 | Positive lead screening | 23 | 2,054 |
| Multiple sclerosis | 1 | 120 | Other gastrointestinal condition (1) | 47 | 2024 | Seizures | 117 | 1,970 |
| Psoriasis | 20 | 122 | Other gastrointestinal condition (2) | 2 | 2090 | Stroke | 0 | 144 |
| Rheumatoid arthritis (adult) | 1 | 141 | | | | Tic's/Tourette's | 51 | 1,961 |
| Rheumatoid arthritis (juvenile) | 1 | 118 | | | | Tuberous sclerosis | 1 | 2,085 |
| Systemic lupus erythematosus | 1 | 121 | | | | | | |
| Other autoimmune condition | 26 | 129 | | | | | | |

[a]In the case of “CT/MRI done,” "EEG done," and "PET done," only participants who had an atypical scan were marked as having at least one condition in the larger category.

[b]Being "excessively clumsy/uncoordinated" was only considered to be a neurological condition if the participant was subsequently diagnosed with a neurological condition.

**Supplementary Table 4**

*Extensive goodness-of-fit criteria for two- through six-class models estimated using latent class analysis.*

| Model | *df* | Gsq | Llik | AIC | AICc | cAIC | mAIC | BIC | aBIC | HQ | HT | Entropy | Smallest Class (%) |
|---|---|---|---|---|---|---|---|---|---|---|---|---|---|
| 2-class | 18 | 189.39 | -5791.64 | 11609.27 | 11609.44 | 11695.68 | 11622.27 | 11682.68 | 11641.38 | 11636.16 | 11609.48 | 3.19 | 37.40 |
| 3-class | 10 | 65.95 | -5726.88 | 11495.77 | 11496.20 | 11635.34 | 11516.77 | 11614.34 | 11547.62 | 11539.21 | 11496.26 | 3.14 | 22.19 |
| 4-class | 2 | 10.28 | -5691.07 | 11440.13 | 11440.95 | 11632.88 | 11469.13 | 11603.88 | 11511.74 | 11500.12 | 11441.04 | 3.12 | 14.74 |
| 5-class | -6 | 7.55 | -5685.83 | 11445.66 | 11446.99 | 11691.58 | 11482.66 | 11654.58 | 11537.02 | 11522.19 | 11447.10 | 3.12 | 2.40 |
| 6-class | -14 | 189.39 | -5791.64 | 11673.27 | 11675.25 | 11972.36 | 11718.27 | 11927.36 | 11784.39 | 11766.36 | 11675.39 | 3.18 | 1.29 |

*Note.* Latent class analysis was conducted with five *DSM*-Oriented Scales of the Child Behavior Checklist for Ages 6 to 18 (Anxiety Problems, Affective Problems, Attention Deficit/Hyperactivity Problems, Conduct Problems, and Oppositional Defiant Problems) as indicator variables. Age and sex were used as covariates.

**Supplementary Figure 2**

*Extensive goodness-of-fit criteria for two- through six-class models estimated using latent class analysis.*

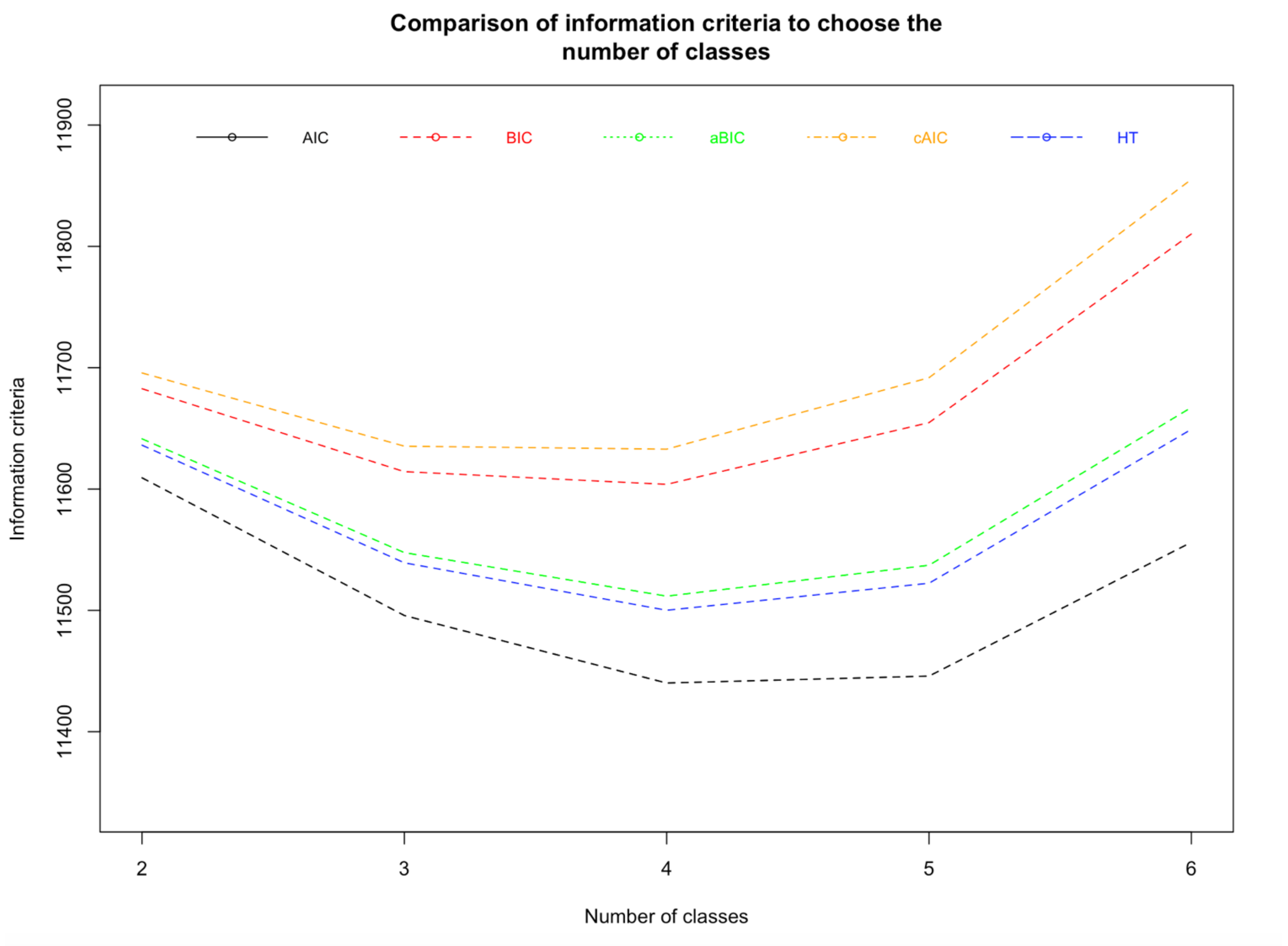

**Supplementary Figure 3**

*Goodness-of-fit criteria for a randomly selected subset of 1,000 participants from the entire latent class analysis sample (n = 2,087) with 1,000 estimations of one to six latent classes.*

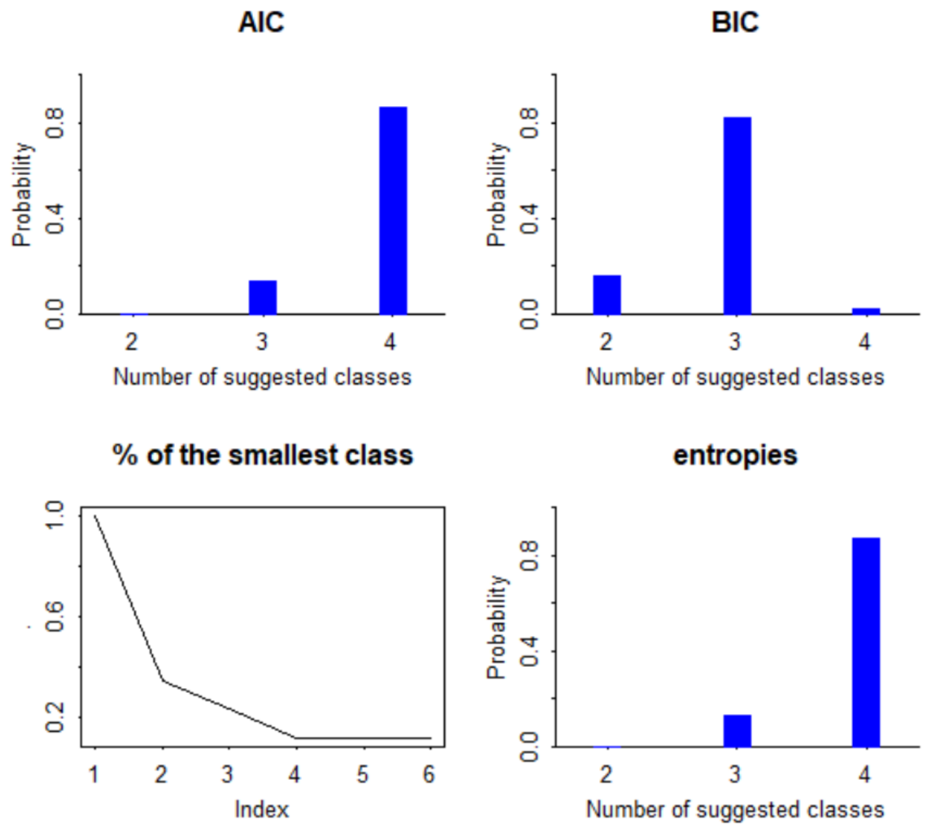

**Supplementary Figure 4**

*Percent of participants in each latent class with borderline/clinical levels of co-occurring psychiatric symptoms.*

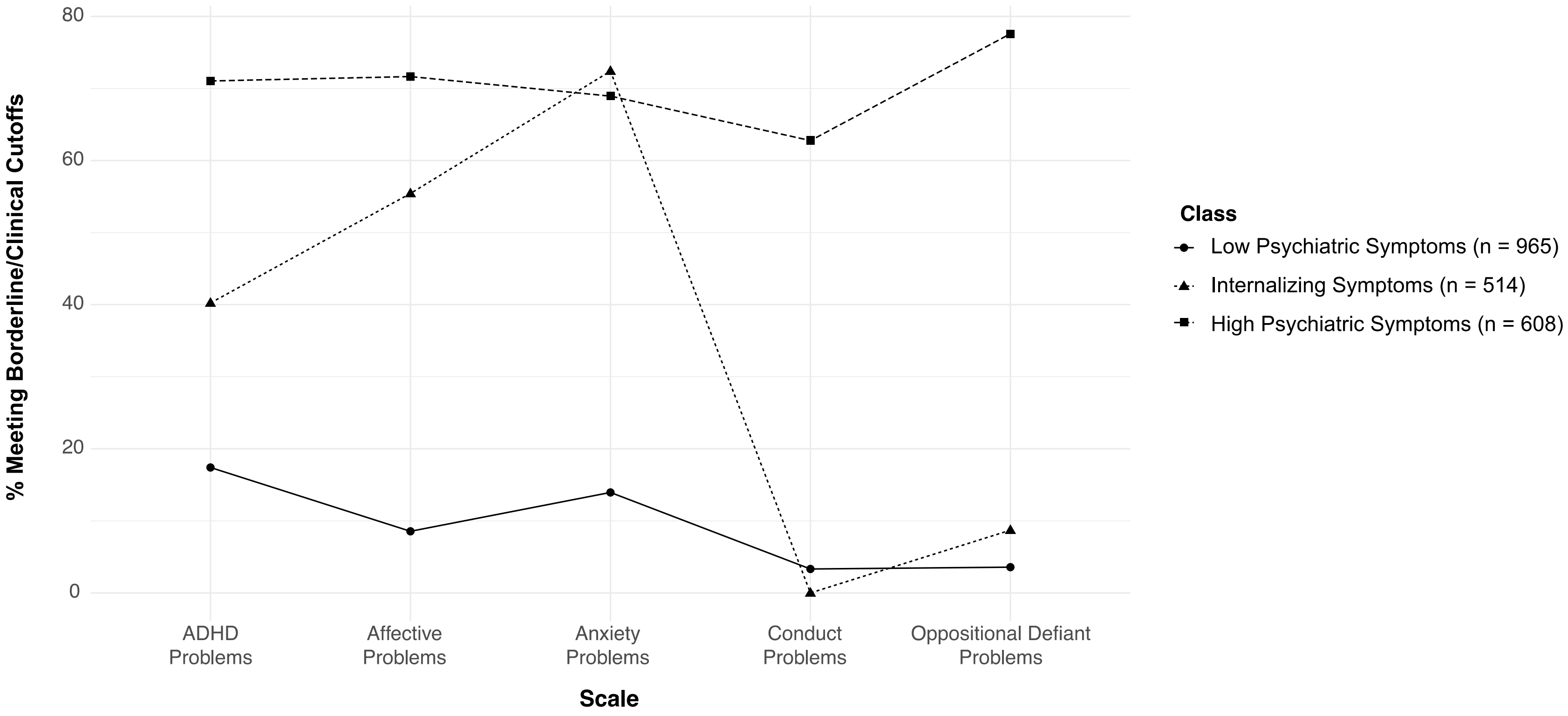

*Note.* Latent class analysis was conducted with five *DSM*-Oriented Scales of the Child Behavior Checklist for Ages 6 to 18 (Anxiety Problems, Affective Problems, Attention Deficit/Hyperactivity Problems, Conduct Problems, and Oppositional Defiant Problems) as indicator variables. Age and sex were used as covariates.

**Supplementary Table 5**

*Percent of participants whose caregiver(s) endorsed a history of each medical condition grouped into one of three larger categories: Allergies/Autoimmune Conditions.*

| Subgroup | Asthma | Bowel Disorders | Hypothyroidism | Other Autoimmune Condition | Psoriasis | Environment Allergies | Food Allergies | Medication Allergies |
|---|---|---|---|---|---|---|---|---|
| Externalizing Symptoms | 6.20 | 4.26 | 0.39 | 1.55 | 2.33 | 24.81 | 15.12 | 11.63 |
| Internalizing Symptoms | 11.68 | 1.82 | 0.15 | 0.91 | 1.37 | 28.98 | 12.75 | 16.54 |
| High Symptoms | 12.10 | 3.18 | 1.27 | 1.59 | 0.32 | 29.94 | 15.29 | 16.56 |

*Note. Only allergies/autoimmune conditions with endorsement rates ≥1% are shown.*

**Supplementary Table 6**

*Percent of participants whose caregiver(s) endorsed a history of each medical condition grouped into one of three larger categories: Gastrointestinal Conditions.*

| Subgroup | Bloating | Constipation | Diarrhea | Excessive Gas | Gastroesophageal Reflux | Irritable Bowel syndrome | Other Gastrointestinal Condition (1) | Unusual Stools | Vomiting |
|---|---|---|---|---|---|---|---|---|---|
| Externalizing Symptoms | 2.58 | 19.77 | 5.43 | 2.33 | 5.04 | 1.16 | 3.49 | 8.14 | 3.49 |
| Internalizing Symptoms | 2.58 | 21.24 | 7.74 | 3.49 | 7.44 | 0.46 | 2.28 | 5.31 | 4.10 |
| High Symptoms | 3.18 | 19.75 | 9.24 | 5.41 | 9.24 | 0.32 | 2.23 | 5.10 | 4.78 |

*Note. Only gastrointestinal conditions with endorsement rates ≥1% are shown.*

**Supplementary Table 7**

*Percent of participants whose caregiver(s) endorsed a history of each medical condition grouped into one of three larger categories: Neurological Conditions.*

| Subgroup | EEG Done | Excessively Clumsy/Uncoordinated | Meningitis | Movement Abnormalities | MRI/CT Done | Positive Lead Screening | Tourette's/ Tics | Febrile Seizures | Seizures Other |
|---|---|---|---|---|---|---|---|---|---|
| Externalizing Symptoms | 3.10 | 8.53 | 1.16 | 2.33 | 2.71 | 0.39 | 3.10 | 3.49 | 5.04 |
| Internalizing Symptoms | 4.55 | 10.17 | 0.15 | 1.97 | 1.82 | 1.37 | 12.75 | 3.95 | 6.83 |
| High Symptoms | 3.50 | 11.46 | 0.32 | 0.64 | 1.59 | 1.27 | 15.29 | 3.82 | 5.10 |

*Note.* *Only neurological conditions with endorsement rates ≥1% are shown.*

**Supplementary Table 8**

*Manually-recoded sleep duration (sleep_duration_curr_clean) variable from the Simons Simplex Collection.*

| child_regularity_duration_hours_curr | sleep_duration_curr_clean |
|---|---|
| 7.5 | 7.5 |
| 9 | 9 |
| 10 | 10 |
| 9 | 9 |
| 9 | 9 |
| 10 | 10 |
| About 9 hours | 9 |
| 10 | 10 |
| 9 | 9 |
| 10 | 10 |
| 9 | 9 |
| 9 | 9 |
| 12 | 12 |
| 10 | 10 |
| 8 | 8 |
| 7 | 7 |
| 8 | 8 |
| 12-Nov | 11.5 |
| 11-Oct | 10.5 |
| 8.5 | 8.5 |
| 9 | 9 |
| About 10 hours | 10 |
| 10 | 10 |

| | |
|---|---|
| 11 | 11 |
| 6 | 6 |
| 10.5 | 10.5 |
| 8 | 8 |
| 9 | 9 |
| 8-9 hours | 8.5 |
| 11 | 11 |
| 11 | 11 |
| 10 | 10 |
| 10-Jul | 8.5 |
| 10-Sep | 9.5 |
| 7.5 | 7.5 |
| 12-Nov | 11.5 |
| 9-Aug | 8.5 |
| 11 | 11 |
| 9.5 hrs during the week | 9.5 |
| 8 | 8 |
| 11 | 11 |
| 9 | 9 |
| 8 | 8 |
| 9 | 9 |
| 9 | 9 |
| 10 | 10 |
| 8 | 8 |
| 10 | 10 |
| 10 hours | 10 |
| 10 | 10 |
| 10 | 10 |

| | |
|---|---|
| 10 | 10 |
| 9 | 9 |
| 8 1/2 hours | 8.5 |
| 9 | 9 |
| 8-Jul | 7.5 |
| 10-Sep | 9.5 |
| 11 | 11 |
| 10 | 10 |
| 8 hours | 8 |
| 8 | 8 |
| 8-Jul | 7.5 |
| 8 | 8 |
| 11 | 11 |
| 10.5 | 10.5 |
| 10 | 10 |
| 10.5 | 10.5 |
| 8.5 | 8.5 |
| 10 | 10 |
| 11 | 11 |
| 9 hours | 9 |
| 7 | 7 |
| 9 | 9 |
| 9 | 9 |
| 10hrs | 10 |
| 9 | 9 |
| 11 | 11 |
| 10 | 10 |
| 9 | 9 |

| | |
|---|---|
| 10 | 10 |
| 11-Oct | 10.5 |
| 9 | 9 |
| 5 | 5 |
| 10 | 10 |
| 9.5 | 9.5 |
| 8 | 8 |
| 8 hours | 8 |
| 7.5 hrs | 7.5 |
| 8 | 8 |
| 8 | 8 |
| 9 | 9 |
| 9.5 | 9.5 |
| ~7 | 7 |
| 10-Sep | 9.5 |
| 10 | 10 |
| 8-10 hrs | 9 |
| 10 hours | 10 |
| 9 | 9 |
| 10 | 10 |
| 10 | 10 |
| 10.5 hours | 10.5 |
| 11 | 11 |
| 8 | 8 |
| 8 | 8 |
| 11 | 11 |
| 10 | 10 |
| 7 | 7 |

| | |
|---|---|
| 8-9.5 | 8.75 |
| 9 | 9 |
| 6 | 6 |
| 10 | 10 |
| 9 | 9 |
| 10 | 10 |
| 8.5 | 8.5 |
| 9 | 9 |
| 7 | 7 |
| 7.5-9 hours | 8.25 |
| 10 | 10 |
| 10 | 10 |
| 9 hours | 9 |
| 9-Aug | 8.5 |
| 9-10 hrs | 9.5 |
| 8 | 8 |
| 10-Sep | 9.5 |
| 10-Sep | 9.5 |
| 9.5 | 9.5 |
| 10 | 10 |
| 9-Aug | 8.5 |
| 8 | 8 |
| 8 | 8 |
| 9 | 9 |
| 10 | 10 |
| 10 | 10 |
| 9 | 9 |
| 10 | 10 |

| | |
|---|---|
| 9 | 9 |
| 8 | 8 |
| 8 | 8 |
| 9.5 | 9.5 |
| 10 | 10 |
| 9 | 9 |
| 8 | 8 |
| 9 | 9 |
| 9 | 9 |
| 9 | 9 |
| 9.5 | 9.5 |
| 10 | 10 |
| 10 | 10 |
| 8-Jul | 7.5 |
| 8 | 8 |
| 7-Jun | 6.5 |
| 8 | 8 |
| 8 hrs | 8 |
| 9.75 | 9.75 |
| 10 | 10 |
| 10 | 10 |
| 10-Aug | 9 |
| 9 | 9 |
| 9 | 9 |
| 9.5 | 9.5 |
| 8.5 | 8.5 |

| | |
|---|---|
| 9-10 hours | 9.5 |
| 9 | 9 |
| 9 | 9 |
| 9 | 9 |
| 8-Jul | 7.5 |
| 10 | 10 |
| 11 | 11 |
| 10-Aug | 9 |
| 9 | 9 |
| 7 hrs | 7 |
| 10 | 10 |
| 9 | 9 |
| about 8 hours | 8 |
| 9 | 9 |
| 10 | 10 |
| 9 | 9 |
| 9 hours | 9 |
| 11-Oct | 10.5 |
| 10 | 10 |
| 10-11 hours | 10.5 |
| 10 | 10 |
| 10 | 10 |
| 9 hours | 9 |
| 11 | 11 |
| 10 | 10 |
| 9 hrs | 9 |

| | |
|---|---|
| 9.5 | 9.5 |
| 8 | 8 |
| 8.5 | 8.5 |
| 10 | 10 |
| 9 | 9 |
| 11 | 11 |
| 9 | 9 |
| 8.5 | 8.5 |
| 8 | 8 |
| 9.5 | 9.5 |
| 9 | 9 |
| 9 | 9 |
| 10.5 | 10.5 |
| 7 | 7 |
| 10 | 10 |
| 8 | 8 |
| 9 | 9 |
| 9 | 9 |
| 9.5 | 9.5 |
| 9 | 9 |
| 9 hours | 9 |
| 9.5 | 9.5 |
| 7.5-8 hours | 7.75 |
| 11-Oct | 10.5 |
| 10 | 10 |
| 8 | 8 |
| 10 | 10 |
| 9 | 9 |

| | |
|---|---|
| 8 | 8 |
| don't know | |
| 8 | 8 |
| 8.5 hrs | 8.5 |
| 10 | 10 |
| 10 | 10 |
| 8-9 hours | 8.5 |
| 5-Apr | 4.5 |
| 10 | 10 |
| 9 | 9 |
| 12-Nov | 11.5 |
| 10 | 10 |
| 9.5 | 9.5 |
| 10 | 10 |
| 9 | 9 |
| 10 | 10 |
| 10 | 10 |
| 10 hrs | 10 |
| 9 | 9 |
| 9-10 hrs | 9.5 |
| 8-9 hours | 8.5 |
| 8.5 | 8.5 |
| 8-Jul | 7.5 |
| 10 | 10 |
| 9 | 9 |
| 10 | 10 |
| 9.5 | 9.5 |
| 8 | 8 |

| | |
|---|---|
| 9 | 9 |
| 9-Aug | 8.5 |
| 9 | 9 |
| 10 hrs | 10 |
| 10-Sep | 9.5 |
| 10-Sep | 9.5 |
| 10 hours | 10 |
| 8 hours | 8 |
| 10 | 10 |
| 11 | 11 |
| 9.5 | 9.5 |
| 9 | 9 |
| 9 | 9 |
| 8.5 | 8.5 |
| 10 | 10 |
| 9.5 | 9.5 |
| 11 | 11 |
| 9.5 | 9.5 |
| 8 | 8 |
| 10 | 10 |
| 11 | 11 |
| 10 | 10 |
| 11-Oct | 10.5 |
| 9 | 9 |
| 9.5 | 9.5 |
| 9 | 9 |
| 10 | 10 |
| 8 hours | 8 |

| | |
|---|---|
| 8 | 8 |
| 9 | 9 |
| 11 hours | 11 |
| 10.5 | 10.5 |
| 10 | 10 |
| 9 | 9 |
| 8 | 8 |
| 9 | 9 |
| 10 hours | 10 |
| 11-Oct | 10.5 |
| 10 | 10 |
| 8 | 8 |
| 10-Sep | 9.5 |
| 10.5 | 10.5 |
| 10 | 10 |
| 10 | 10 |
| 8 | 8 |
| 11 | 11 |
| 10 | 10 |
| 10 | 10 |
| 10.5 | 10.5 |
| 8 | 8 |
| 10 | 10 |
| 9 hours | 9 |
| 11 hours | 11 |
| 10 | 10 |
| 8 | 8 |
| 8 | 8 |

| | |
|---|---|
| 9 hours | 9 |
| 8 | 8 |
| 9 | 9 |
| 9 | 9 |
| 10.5 hours | 10.5 |
| 10 | 10 |
| 8.5 | 8.5 |
| 10 | 10 |
| 7 | 7 |
| 11-Oct | 10.5 |
| 10 hours | 10 |
| 10 | 10 |
| 11 | 11 |
| 7-Jun | 6.5 |
| 11.5 | 11.5 |
| 9 | 9 |
| 9 | 9 |
| 10.5 | 10.5 |
| 11 hrs | 11 |
| 13 | 13 |
| 10 | 10 |
| 4 | 4 |
| 8.5 | 8.5 |
| 10 | 10 |
| 10 hours | 10 |
| 7.5 | 7.5 |
| 10 | 10 |

| | |
|---|---|
| 8 hours | 8 |
| 9.5 | 9.5 |
| 10 | 10 |
| 8hrs | 8 |
| 10.5 | 10.5 |
| 9 | 9 |
| 10-Sep | 9.5 |
| 8 | 8 |
| 7 | 7 |
| 10 | 10 |
| 8 | 8 |
| 9 | 9 |
| 7.5 | 7.5 |
| 8 | 8 |
| 8-Jul | 7.5 |
| 10 | 10 |
| 11-Oct | 10.5 |
| 10.5 | 10.5 |
| 10 | 10 |
| 9-Aug | 8.5 |
| 9.5 | 9.5 |
| 10 | 10 |
| 10 hrs | 10 |
| 9 | 9 |
| 8 hours | 8 |
| 7.5 | 7.5 |
| 9 | 9 |
| 8 | 8 |

| | |
|---|---|
| 11 | 11 |
| 8 | 8 |
| 10 | 10 |
| 91/2-10 hours | 9.75 |
| 10 | 10 |
| 8 hrs | 8 |
| 9 | 9 |
| 12-Oct | 11 |
| 11 | 11 |
| 9.5 | 9.5 |
| 10 | 10 |
| 10 | 10 |
| 8 | 8 |
| 11 | 11 |
| 7.5 | 7.5 |
| 7 | 7 |
| 11 | 11 |
| 9 | 9 |
| 9 | 9 |
| 8 | 8 |
| 9 | 9 |
| 8 | 8 |
| 8.5hrs | 8.5 |
| 6-Sep | 7.5 |
| 8.5 | 8.5 |
| 10-Aug | 9 |
| 10 | 10 |

| | |
|---|---|
| 6 | 6 |
| 8.5 | 8.5 |
| 10 | 10 |
| 10 | 10 |
| 7 hours | 7 |
| 10 | 10 |
| 9 | 9 |
| 10 | 10 |
| 6 | 6 |
| 9 | 9 |
| 7 | 7 |
| 9-Aug | 8.5 |
| 7.5 | 7.5 |
| 8-9 hours | 8.5 |
| 11 hours | 11 |
| 8 | 8 |
| 7 | 7 |
| 9 | 9 |
| 6.5 HRS | 6.5 |
| 9 | 9 |
| 9 hrs | 9 |
| 9 | 9 |
| 9 hrs | 9 |
| 10 | 10 |
| 10-12 hrs | 11 |
| 10 | 10 |
| 10.5 | 10.5 |
| 9 | 9 |

| | |
|---|---|
| 8 | 8 |
| 5.5 | 5.5 |
| 11-Sep | 10 |
| 11 | 11 |
| 10 | 10 |
| 10-Sep | 9.5 |
| 10.5 | 10.5 |
| 9 | 9 |
| 9 | 9 |
| 8 hrs | 8 |
| 10 | 10 |
| 9 | 9 |
| 8 | 8 |
| normal | |
| 8 hours | 8 |
| 10 | 10 |
| 7 | 7 |
| 9-Aug | 8.5 |
| 9.5 | 9.5 |
| 10 | 10 |
| 9 | 9 |
| 8 | 8 |
| 7 | 7 |
| 8.5 | 8.5 |
| 12 | 12 |
| 8 | 8 |
| 10.5 | 10.5 |
| 9 | 9 |

| | |
|---|---|
| 8 | 8 |
| 10 | 10 |
| 6 | 6 |
| 10 | 10 |
| 9 | 9 |
| 8.5 | 8.5 |
| 10-Aug | 9 |
| 7 | 7 |
| 9 | 9 |
| 11 | 11 |
| 10 | 10 |
| 9 | 9 |
| 11-Oct | 10.5 |
| 10 | 10 |
| 6.5 | 6.5 |
| 8 | 8 |
| 8.5 | 8.5 |
| 9 | 9 |
| 8 | 8 |
| 9 | 9 |
| 10 | 10 |
| 7.5 | 7.5 |
| 10 | 10 |
| 12 | 12 |
| 8 | 8 |
| 6 | 6 |
| 7 | 7 |
| 11 | 11 |

| | |
|---|---|
| 9 | 9 |
| 10 | 10 |
| 9 | 9 |
| 9 | 9 |
| 9 | 9 |
| 10.5 | 10.5 |
| 8 | 8 |
| 8 | 8 |
| 10 | 10 |
| 10 | 10 |
| 9 | 9 |
| 10.5 | 10.5 |
| 10 | 10 |
| 7 | 7 |
| 10 | 10 |
| 7.5 | 7.5 |
| 8 | 8 |
| 10 | 10 |
| 9 | 9 |
| 10-Sep | 9.5 |
| 10 | 10 |
| 10 | 10 |
| 9 | 9 |
| 8 | 8 |
| 10 | 10 |
| 10 | 10 |
| 12 | 12 |
| 10-Sep | 9.5 |

| | |
|---|---|
| 12 | 12 |
| 9 | 9 |
| 9 | 9 |
| 8 | 8 |
| 8 | 8 |
| 12 | 12 |
| 10 | 10 |
| 9 | 9 |
| 11 | 11 |
| 9 | 9 |
| 10 | 10 |
| 10 | 10 |
| 8 | 8 |
| 8 | 8 |
| 8 | 8 |
| 10 | 10 |
| 8 | 8 |
| 9 | 9 |
| 7.5 | 7.5 |
| 8-Jul | 7.5 |
| 12 | 12 |
| 12-Oct | 11 |
| 9 | 9 |
| 9 | 9 |
| 9.5 | 9.5 |
| 8 | 8 |
| 9 | 9 |
| 9 | 9 |

| | |
|---|---|
| 10 | 10 |
| 10 | 10 |
| 10.5 | 10.5 |
| 8 | 8 |
| 9.5 | 9.5 |
| 8 | 8 |
| 9.5 | 9.5 |
| 9 | 9 |
| 6 | 6 |
| 8 | 8 |
| 10 | 10 |
| 8 | 8 |
| 8 hrs | 8 |
| 10 | 10 |
| 8 | 8 |
| 11 | 11 |
| 8 | 8 |
| 8 | 8 |
| 10.5 | 10.5 |
| 9 | 9 |
| 10 | 10 |
| 8 | 8 |
| 10 | 10 |
| 10 | 10 |
| 9 | 9 |
| 7 | 7 |
| 8-Jul | 7.5 |

| | |
|---|---|
| 10 | 10 |
| 8 | 8 |
| 10 | 10 |
| 11.5 | 11.5 |
| 10 | 10 |
| 11 | 11 |
| 9 | 9 |
| 9.5 | 9.5 |
| 11 | 11 |
| 8 | 8 |
| 10 | 10 |
| 7 | 7 |
| 7 | 7 |
| 9 | 9 |
| 9 hrs | 9 |
| 8 | 8 |
| 10 | 10 |
| 9.5 | 9.5 |
| 11 | 11 |
| 9.5 | 9.5 |
| 9 hrs | 9 |
| 9 | 9 |
| 11 | 11 |
| 10 | 10 |
| 5 | 5 |
| 8 | 8 |
| 10 | 10 |
| 9 | 9 |

| | |
|---|---|
| 10.5 | 10.5 |
| 9 | 9 |
| 10 | 10 |
| 10 | 10 |
| 9-Aug | 8.5 |
| 11 | 11 |
| 9.5 | 9.5 |
| 6 | 6 |
| 10-Jan | 5.5 |
| 10 | 10 |
| 9.5 | 9.5 |
| 11 | 11 |
| 9 | 9 |
| 9.5 | 9.5 |
| 10 | 10 |
| 10 | 10 |
| 9 | 9 |
| 8 | 8 |
| 11 | 11 |
| 9 | 9 |
| 9 | 9 |
| 10 | 10 |
| 8 | 8 |
| 10 | 10 |
| 9.5 | 9.5 |
| 9 | 9 |
| 10.25 | 10.25 |
| 9 | 9 |

| | |
|---|---|
| 9.5 | 9.5 |
| 9 | 9 |
| 10 | 10 |
| 10 | 10 |
| 9 | 9 |
| 8 | 8 |
| 9 | 9 |
| 8 | 8 |
| 6 | 6 |
| 9 | 9 |
| 10 | 10 |
| 11 | 11 |
| 9.5 | 9.5 |
| 10 | 10 |
| 8 | 8 |
| 9 | 9 |
| 10 | 10 |
| 12-Oct | 11 |
| 9 | 9 |
| 9 | 9 |
| 8 | 8 |
| 10 | 10 |
| 10.5 | 10.5 |
| 10.5 | 10.5 |
| 9 | 9 |
| 8.5 | 8.5 |
| 9 | 9 |
| 8 | 8 |

| | |
|---|---|
| 10 | 10 |
| 10.5 | 10.5 |
| 11 | 11 |
| 8 | 8 |
| 10 | 10 |
| 8 | 8 |
| 9 | 9 |
| 10 | 10 |
| 8 | 8 |
| 9.5 | 9.5 |
| 9 | 9 |
| 9 | 9 |
| 9 | 9 |
| 10 | 10 |
| 10 | 10 |
| 8 | 8 |
| 11 | 11 |
| 9.5 | 9.5 |
| 9 | 9 |
| 8 | 8 |
| 9 | 9 |
| 9 | 9 |
| 9 | 9 |
| 10 | 10 |
| 10 | 10 |
| 9.5 | 9.5 |
| 10 | 10 |
| 9-Aug | 8.5 |

| | |
|---|---|
| 7 | 7 |
| 10 | 10 |
| 9 | 9 |
| 10 | 10 |
| 10 | 10 |
| 9 | 9 |
| 9.5 | 9.5 |
| 6 | 6 |
| 10.5 | 10.5 |
| 8.5 | 8.5 |
| 8.5 | 8.5 |
| 10 | 10 |
| 9 | 9 |
| 11 | 11 |
| 10 | 10 |
| 7 | 7 |
| 8hrs | 8 |
| 10 | 10 |
| 11 | 11 |
| 10 | 10 |
| 10 | 10 |
| 9 | 9 |
| 10 | 10 |
| 10 | 10 |
| 7.5 | 7.5 |
| 9 | 9 |
| 9 | 9 |
| 10 | 10 |

| | |
|---|---|
| 11 | 11 |
| 7.5 | 7.5 |
| 9.5 | 9.5 |
| 5 | 5 |
| 9 | 9 |
| 9.5 | 9.5 |
| 10.5 | 10.5 |
| 9 | 9 |
| 10.5 | 10.5 |
| 10 | 10 |
| 9 | 9 |
| 10 | 10 |
| 10.5 | 10.5 |
| 8 | 8 |
| 10.5 | 10.5 |
| 9 | 9 |
| 10 | 10 |
| 10 | 10 |
| 9 | 9 |
| 12 | 12 |
| 10.5 | 10.5 |
| 10 | 10 |
| 11 | 11 |
| 11 | 11 |
| 8 | 8 |
| 8 | 8 |
| 8 | 8 |
| 6 | 6 |

| | |
|---|---|
| 8 | 8 |
| 7.5 | 7.5 |
| 9 | 9 |
| 9.5 | 9.5 |
| 10 | 10 |
| 10 | 10 |
| 10 | 10 |
| 8 | 8 |
| 8 | 8 |
| 12 | 12 |
| 11 | 11 |
| 10.5 | 10.5 |
| 8.5 | 8.5 |
| 9 | 9 |
| 11 | 11 |
| 7 | 7 |
| 10 | 10 |
| 8 | 8 |
| 10 | 10 |
| 10.5 | 10.5 |
| 10.5 | 10.5 |
| 8 | 8 |
| 10 | 10 |
| 10.5 | 10.5 |
| 8 | 8 |
| 9.5 | 9.5 |
| 9.5 | 9.5 |
| 8 | 8 |

| | |
|---|---|
| 10 | 10 |
| 11 | 11 |
| 9.5 | 9.5 |
| 9.5 | 9.5 |
| 10 | 10 |
| 11 | 11 |
| 10 | 10 |
| 9 | 9 |
| 10 | 10 |
| 7 | 7 |
| 8 | 8 |
| 10.5 | 10.5 |
| 8 | 8 |
| 8 | 8 |
| 8.5 | 8.5 |
| 9 | 9 |
| 9 | 9 |
| 12 | 12 |
| 9 | 9 |
| 9.5 | 9.5 |
| 8 | 8 |
| 6 | 6 |
| 10.5 | 10.5 |
| 10 | 10 |
| 9.5 | 9.5 |
| 9.5 | 9.5 |
| 10 | 10 |
| 9.5 | 9.5 |

| | |
|---|---|
| 8.5 | 8.5 |
| 9 | 9 |
| 12 | 12 |
| 11.5 | 11.5 |
| 10.5 | 10.5 |
| 10 | 10 |
| 9 | 9 |
| 10 | 10 |
| 10 | 10 |
| 11 | 11 |
| 7.5 | 7.5 |
| 9 | 9 |
| 9 | 9 |
| 9 | 9 |
| 10 | 10 |
| 11 | 11 |
| 11 | 11 |
| 9 | 9 |
| 11 | 11 |
| 8 | 8 |
| 10 | 10 |
| 10 | 10 |
| 8.5 | 8.5 |
| 9 | 9 |
| 10 | 10 |
| 8.5 | 8.5 |
| 9 | 9 |
| 11 | 11 |

| | |
|---|---|
| 11 | 11 |
| 9 | 9 |
| 10.5 | 10.5 |
| 10 | 10 |
| 9 | 9 |
| 11 | 11 |
| 6.5 | 6.5 |
| 8 | 8 |
| 6 | 6 |
| 8 | 8 |
| 9 | 9 |
| 10.5 | 10.5 |
| 10.5 | 10.5 |
| 9 | 9 |
| 8 | 8 |
| 10 | 10 |
| 8 | 8 |
| 11 | 11 |
| 10 | 10 |
| 10 | 10 |
| 8.5 | 8.5 |
| 8 | 8 |
| 10 | 10 |
| 10 | 10 |
| 11 | 11 |
| 9 | 9 |
| 10 | 10 |
| 9.75 | 9.75 |

| | |
|---|---|
| 10 | 10 |
| 8 | 8 |
| 10 | 10 |
| 10 | 10 |
| 8 | 8 |
| 10 | 10 |
| 9 | 9 |
| 8 | 8 |
| 10 | 10 |
| 8 | 8 |
| 10 | 10 |
| 8 | 8 |
| 10 | 10 |
| 12 | 12 |
| 8.5 | 8.5 |
| 9 | 9 |
| 7 | 7 |
| 9 | 9 |
| 9 | 9 |
| 8.5 | 8.5 |
| 9.5 | 9.5 |
| 9 | 9 |
| 9 | 9 |
| 10.5 | 10.5 |
| 9 | 9 |
| 9 | 9 |
| 9.5 | 9.5 |
| 10.5 | 10.5 |

| | |
|---|---|
| 9 | 9 |
| 10 | 10 |
| 10 | 10 |
| 9.5 | 9.5 |
| 6 | 6 |
| 10 | 10 |
| 9 | 9 |
| 8 | 8 |
| 7.5 | 7.5 |
| 11 | 11 |
| 11 | 11 |
| 10 | 10 |
| 8 | 8 |
| 9 | 9 |
| 8 | 8 |
| 10.5 | 10.5 |
| 8 | 8 |
| 10 | 10 |
| 8 | 8 |
| 11.5 | 11.5 |
| 9 | 9 |
| 8.5 | 8.5 |
| 10 | 10 |
| 10 | 10 |
| 10.5 | 10.5 |
| 9.5 | 9.5 |
| 10 | 10 |
| 9 | 9 |

| | |
|---|---|
| 11 | 11 |
| 10 | 10 |
| 8 | 8 |
| 10 | 10 |
| 8 | 8 |
| 10 | 10 |
| 8 | 8 |
| 10.5 | 10.5 |
| 9 | 9 |
| 8.5 | 8.5 |
| 8 | 8 |
| 10 | 10 |
| 8 | 8 |
| 10 | 10 |
| 9 | 9 |
| 11.5 | 11.5 |
| 9 | 9 |
| 9 | 9 |
| 9 | 9 |
| 10 | 10 |
| 9 | 9 |
| 9 | 9 |
| 12 | 12 |
| 8.5 | 8.5 |
| 9 | 9 |
| 9 | 9 |
| 9 | 9 |
| 10 | 10 |

| | |
|---|---|
| 8 | 8 |
| 9 | 9 |
| 10 | 10 |
| 11 | 11 |
| 8 | 8 |
| 7 | 7 |
| 10.5 | 10.5 |
| 10 | 10 |
| 9 | 9 |
| 10 | 10 |
| 9.5 | 9.5 |
| 10 | 10 |
| 6.5 | 6.5 |
| 8 | 8 |
| 9 | 9 |
| 11 | 11 |
| 7.5 | 7.5 |
| 8 | 8 |
| 7.5 | 7.5 |
| 10 | 10 |
| 10 | 10 |
| 10 | 10 |
| 9.5 | 9.5 |
| 11 | 11 |
| 8 | 8 |
| 9.5 | 9.5 |
| 9 | 9 |
| 9.5 | 9.5 |

| | |
|---|---|
| 11 | 11 |
| 10 | 10 |
| 8 | 8 |
| 8 | 8 |
| 7 | 7 |
| 11 | 11 |
| 10 | 10 |
| 9 | 9 |
| 9.5 | 9.5 |
| 10 | 10 |
| 8 | 8 |
| 8 | 8 |
| 5.5 | 5.5 |
| 10 | 10 |
| 10 | 10 |
| 7 | 7 |
| 9 | 9 |
| 8.5 | 8.5 |
| 9 | 9 |
| 10 | 10 |
| 7 | 7 |
| 9.5 | 9.5 |
| 6.5 | 6.5 |
| 10 | 10 |
| 8.5 | 8.5 |
| 8.5 | 8.5 |
| 8 | 8 |

| | |
|---|---|
| 8.5 | 8.5 |
| 9 | 9 |
| 9 | 9 |
| 7.5 | 7.5 |
| 11 | 11 |
| 9 | 9 |
| 11 | 11 |
| 11 | 11 |
| 9.5 | 9.5 |
| 9 | 9 |
| 8 | 8 |
| 9 | 9 |
| 10 | 10 |
| 7.5 | 7.5 |
| 8 | 8 |
| 10 | 10 |
| 9 | 9 |
| 6 | 6 |
| 9 | 9 |
| 11 | 11 |
| 9.5 | 9.5 |
| 7 | 7 |
| 9.5 | 9.5 |
| 9 | 9 |
| 8 | 8 |
| 8.5 | 8.5 |
| 8.5 | 8.5 |
| 8.5 | 8.5 |

| | |
|---|---|
| 9 | 9 |
| 10 | 10 |
| 4.5 | 4.5 |
| 8 | 8 |
| 10 | 10 |
| 11 | 11 |
| 9 | 9 |
| 9.5 | 9.5 |
| 8 | 8 |
| 10 | 10 |
| 8 | 8 |
| 10 | 10 |
| 9 | 9 |
| 8 | 8 |
| 8 | 8 |
| 10 | 10 |
| 9 | 9 |
| 7.5 | 7.5 |
| 10 | 10 |
| 8 | 8 |
| 9 | 9 |
| 10.5 | 10.5 |
| 8 | 8 |
| 11 | 11 |
| 10 | 10 |
| 10 | 10 |
| 9 | 9 |
| 10 | 10 |

| | |
|---|---|
| 8.5 | 8.5 |
| 11 | 11 |
| 9 | 9 |
| 8 | 8 |
| 9 | 9 |
| 9 | 9 |
| 8 | 8 |
| 10 | 10 |
| 8 | 8 |
| 9 | 9 |
| 10 | 10 |
| 11 | 11 |
| 10 | 10 |
| 9.5 | 9.5 |
| 9.5 | 9.5 |
| 11 | 11 |
| 9.5 | 9.5 |
| 10 | 10 |
| 8 | 8 |
| 12 | 12 |
| 10 | 10 |
| 7 | 7 |
| 8 | 8 |
| 9.5 | 9.5 |
| 11 | 11 |
| 6.5 | 6.5 |
| 10 | 10 |
| 9 | 9 |

| | |
|---|---|
| 11 | 11 |
| 8 | 8 |
| 9 | 9 |
| 9 | 9 |
| 7 | 7 |
| 10 | 10 |
| 9.5 | 9.5 |
| 8 | 8 |
| 10.5 | 10.5 |
| 10.5 | 10.5 |
| 10.5 | 10.5 |
| 8 | 8 |
| 7.5 | 7.5 |
| 9 | 9 |
| 8.5 | 8.5 |
| 10 | 10 |
| 11 | 11 |
| 12 | 12 |
| 9 | 9 |
| 10 | 10 |
| 9 | 9 |
| 10 | 10 |
| 10 | 10 |
| 9 | 9 |
| 8.5 | 8.5 |
| 10 | 10 |
| 10 | 10 |
| 10 | 10 |

| | |
|---|---|
| 10 | 10 |
| 7 | 7 |
| 9 | 9 |
| 10 | 10 |
| 10 | 10 |
| 9 | 9 |
| 9 | 9 |
| 8 | 8 |
| 9 | 9 |
| 9.5 | 9.5 |
| 10 | 10 |
| 9 | 9 |
| 6 | 6 |
| 10 | 10 |
| 10 | 10 |
| 10 | 10 |
| 6 | 6 |
| 10 | 10 |
| 10 | 10 |
| 10 | 10 |
| 10 | 10 |
| 9 | 9 |
| 9 | 9 |
| 10 | 10 |
| 10 | 10 |
| 9 | 9 |
| 7 | 7 |

| | |
|---|---|
| 10 | 10 |
| 6 | 6 |
| 9.5 | 9.5 |
| 9 | 9 |
| 8 | 8 |
| 6 | 6 |
| 11.5 | 11.5 |
| 10 | 10 |
| 10 | 10 |
| 9 | 9 |
| 9.75 | 9.75 |
| 12 | 12 |
| 8 | 8 |
| 10.5 | 10.5 |
| 9 | 9 |
| 11.5 | 11.5 |
| 9 | 9 |
| 10 | 10 |
| 10 | 10 |
| 9 | 9 |
| 10.5 | 10.5 |
| 9 | 9 |
| 9 | 9 |
| 8 | 8 |
| 10 | 10 |
| 10 | 10 |
| 10 | 10 |
| 10 | 10 |

| | |
|---|---|
| 9 | 9 |
| 9 | 9 |
| 8 | 8 |
| 10 | 10 |
| 10.5 | 10.5 |
| 9 | 9 |
| 9 | 9 |
| 11 | 11 |
| 11 | 11 |
| 11 | 11 |
| 11 | 11 |
| 9 | 9 |
| 10 | 10 |
| 10 | 10 |
| 10 | 10 |
| 10 | 10 |
| 9.5 | 9.5 |
| 7 | 7 |
| 10 | 10 |
| 10 | 10 |
| 10.5 | 10.5 |
| 10.5 | 10.5 |
| 10 | 10 |
| 10.5 | 10.5 |
| 9.5 | 9.5 |
| 9 | 9 |
| 7.5 | 7.5 |
| 11.5 | 11.5 |

| | |
|---|---|
| 6.5 | 6.5 |
| 9.5 | 9.5 |
| 9.5 | 9.5 |
| 9 | 9 |
| 10.5 | 10.5 |
| 10 | 10 |
| 8 | 8 |
| 9.5 | 9.5 |
| 9.5 | 9.5 |
| 9 | 9 |
| 10 | 10 |
| 10 | 10 |
| 8.5 | 8.5 |
| 10 | 10 |
| 7 | 7 |
| 8 | 8 |
| 10 | 10 |
| 10 | 10 |
| 9 | 9 |
| 10 | 10 |
| 11 | 11 |
| 12 | 12 |
| 11 | 11 |
| 9.5 | 9.5 |
| 11 | 11 |
| 11 | 11 |
| 11.5 | 11.5 |
| 11 | 11 |

| | |
|---|---|
| 9 | 9 |
| 9 | 9 |
| 10 | 10 |
| 10 | 10 |
| 10 | 10 |
| 8 | 8 |
| 9 | 9 |
| 10 | 10 |
| 10 | 10 |
| 9 | 9 |
| 11 | 11 |
| 10 | 10 |
| 10 | 10 |
| 9 | 9 |
| 9 | 9 |
| 10 | 10 |
| 10 | 10 |
| 9 | 9 |
| 10 | 10 |
| 10 | 10 |
| 8 | 8 |
| 10 | 10 |
| 9 | 9 |
| 10 | 10 |
| 10 | 10 |
| 10 | 10 |
| 11 | 11 |
| 8 | 8 |

| | |
|---|---|
| 11 | 11 |
| 9.5 | 9.5 |
| 9 | 9 |
| 9 | 9 |
| 7.5 | 7.5 |
| 11 | 11 |
| 12 | 12 |
| 11 | 11 |
| 8 | 8 |
| 11 | 11 |
| 6 | 6 |
| 9 | 9 |
| 10 | 10 |
| 9 | 9 |
| 12 | 12 |
| 10 | 10 |
| 10 | 10 |
| 9 | 9 |
| 10 | 10 |
| 8 | 8 |
| 11 | 11 |
| 9 | 9 |
| 11.75 | 11.75 |
| 9 | 9 |
| 9.5 | 9.5 |
| 9 | 9 |
| 10 | 10 |
| 9 | 9 |

| | |
|---|---|
| 9 | 9 |
| 8 | 8 |
| 10 | 10 |
| 11 | 11 |
| 10 | 10 |
| 10 | 10 |
| 8 | 8 |
| 9.5 | 9.5 |
| 10 | 10 |
| 8.5 | 8.5 |
| 9.5 | 9.5 |
| 9 | 9 |
| 10.5 | 10.5 |
| 9 | 9 |
| 8.5 | 8.5 |
| 10 | 10 |
| 9 | 9 |
| 11.5 | 11.5 |
| 10 | 10 |
| 11 | 11 |
| 10 | 10 |
| 10.5 | 10.5 |
| 7.5 | 7.5 |
| 10.5 | 10.5 |
| 11 | 11 |
| 10 | 10 |
| 9 | 9 |
| 10 | 10 |

| | |
|---|---|
| 10 | 10 |
| 10 | 10 |
| 9 | 9 |
| 7 | 7 |
| 8.5 | 8.5 |
| 10 | 10 |
| 9 | 9 |
| 10 | 10 |
| 10 | 10 |
| 10 | 10 |
| 10 | 10 |
| 8 | 8 |
| 9.5 | 9.5 |
| 9 | 9 |
| 8 | 8 |
| 10.5 | 10.5 |
| 9 | 9 |
| 9.5 | 9.5 |
| 8.5 | 8.5 |
| 10 | 10 |
| 9 | 9 |
| 9.5 | 9.5 |
| 9 | 9 |
| 7.5 | 7.5 |
| 6 | 6 |
| 12 | 12 |
| 11 | 11 |
| 10 | 10 |

| | |
|---|---|
| 11 | 11 |
| 7 | 7 |
| 10 | 10 |
| 9.5 | 9.5 |
| 8.5 | 8.5 |
| 9 | 9 |
| 8 | 8 |
| 9.5 | 9.5 |
| 8 | 8 |
| 10 | 10 |
| 6.5 | 6.5 |
| 10 | 10 |
| 11 | 11 |
| 9 | 9 |
| 10 | 10 |
| 9.5 | 9.5 |
| 10.5 | 10.5 |
| 7.5 | 7.5 |
| 8 | 8 |
| 7.5 | 7.5 |
| 10 | 10 |
| 11 | 11 |
| 10 | 10 |
| 13 | 13 |
| 10 | 10 |
| 9 | 9 |
| 9 | 9 |
| 12 | 12 |

| | |
|---|---|
| 7 | 7 |
| 5.5 | 5.5 |
| 8.5 | 8.5 |
| 10.5 | 10.5 |
| 8 | 8 |
| 10 | 10 |
| 11 | 11 |
| 9 | 9 |
| 8 | 8 |
| 8.5 | 8.5 |
| 9.5 | 9.5 |
| 11 | 11 |
| 8 | 8 |
| 10 | 10 |
| 9 | 9 |
| 10.5 | 10.5 |
| 9.5 | 9.5 |
| 8 | 8 |
| 9 | 9 |
| 11 | 11 |
| 10 | 10 |
| 8 | 8 |
| 7 | 7 |
| 9 | 9 |
| 10 | 10 |
| 8.5 | 8.5 |
| 11.5 | 11.5 |

| | |
|---|---|
| 9.5 | 9.5 |
| 10 | 10 |
| 7 | 7 |
| 8 | 8 |
| 8 | 8 |
| 10 | 10 |
| 9 | 9 |
| 9 | 9 |
| 8.5 | 8.5 |
| 9.5 | 9.5 |
| 8 | 8 |
| 10 | 10 |
| 10 | 10 |
| 10 | 10 |
| 8.5 | 8.5 |
| 9 | 9 |
| 9.5 | 9.5 |
| 9 | 9 |
| 8 | 8 |
| 9.5 | 9.5 |
| 10 | 10 |
| 8 | 8 |
| 7.75 | 7.75 |
| 11 | 11 |
| 11 | 11 |
| 8.5 | 8.5 |
| 9 | 9 |
| 8.5 | 8.5 |

| | |
|---|---|
| 9 | 9 |
| 9 | 9 |
| 8 | 8 |
| 7.5 | 7.5 |
| 10 | 10 |
| 9.5 | 9.5 |
| 10 | 10 |
| 10 | 10 |
| 8 | 8 |
| 8.5 | 8.5 |
| 6 | 6 |
| 10 | 10 |
| 8 | 8 |
| 9 | 9 |
| 6 | 6 |
| 9 | 9 |
| 8 | 8 |
| 9.5 | 9.5 |
| 8 | 8 |
| 10 | 10 |
| 10 | 10 |
| 8 | 8 |
| 9 | 9 |
| 5.5 | 5.5 |
| 10 | 10 |
| 9 | 9 |
| 11 | 11 |

| | |
|---|---|
| 9.5 | 9.5 |
| 7.5 | 7.5 |
| 10 | 10 |
| 8 | 8 |
| 8.5 | 8.5 |
| 10 | 10 |
| 10.5 | 10.5 |
| 9.5 | 9.5 |
| 11 | 11 |
| 9.5 | 9.5 |
| 10 | 10 |
| 9 | 9 |
| 10 | 10 |
| 11.5 | 11.5 |
| 12 | 12 |
| 10 | 10 |
| 9 | 9 |
| 7 | 7 |
| 8 | 8 |
| 11 | 11 |
| 8 | 8 |
| 9 | 9 |
| 10 | 10 |
| 8.5 | 8.5 |
| 10.5 | 10.5 |
| 9 | 9 |
| 10.5 | 10.5 |

| | |
|---|---|
| 8 | 8 |
| 10 | 10 |
| 9.5 | 9.5 |
| 10 | 10 |
| 9 | 9 |
| 10 | 10 |
| 10 | 10 |
| 9 | 9 |
| 8 | 8 |
| 9 | 9 |
| 9.5 | 9.5 |
| 8.5 | 8.5 |
| 10 | 10 |
| 8.5 | 8.5 |
| 10 | 10 |
| 11 | 11 |
| 9 | 9 |
| 9 | 9 |
| 10 | 10 |
| 9 | 9 |
| 7 | 7 |
| 10 | 10 |
| 9 | 9 |
| 10 | 10 |
| 8.5 | 8.5 |
| 9.5 | 9.5 |
| 9 | 9 |
| 8.5 | 8.5 |

| | |
|---|---|
| 11 | 11 |
| 9 | 9 |
| 9.5 | 9.5 |
| 9.5 | 9.5 |
| 10 | 10 |
| 10.5 | 10.5 |
| 9 | 9 |
| 11 | 11 |
| 12 | 12 |
| 9 | 9 |
| 10 | 10 |
| 10 | 10 |
| 8 | 8 |
| 8 | 8 |
| 8 | 8 |
| 7 | 7 |
| 8 | 8 |
| 10 | 10 |
| 7 | 7 |
| 9.5 | 9.5 |
| 9 | 9 |
| 8 | 8 |
| 7 | 7 |
| 10 | 10 |
| 8.5 | 8.5 |
| 9 | 9 |
| 9 | 9 |
| 8 | 8 |

| | |
|---|---|
| 9 | 9 |
| 9 | 9 |
| 11 | 11 |
| 9.5 | 9.5 |
| 10 | 10 |
| 10 | 10 |
| 6 | 6 |
| 6 | 6 |
| 8 | 8 |
| 10 | 10 |
| 8 | 8 |
| 9 | 9 |
| 8.5 | 8.5 |
| 10 | 10 |
| 10 | 10 |
| 9 | 9 |
| 9.5 | 9.5 |
| 10 | 10 |
| 10 | 10 |
| 10 | 10 |
| 9 | 9 |
| 10 | 10 |
| 10 | 10 |
| 10 | 10 |
| 10.5 | 10.5 |
| 9.5 | 9.5 |
| 9.5 | 9.5 |
| 9 | 9 |

| | |
|---|---|
| 11 | 11 |
| 7 | 7 |
| 11 | 11 |
| 9 | 9 |
| 9 | 9 |
| 11 | 11 |
| 9 | 9 |
| 10 | 10 |
| 9 | 9 |
| 9 | 9 |
| 9 | 9 |
| 11 | 11 |
| 9 | 9 |
| 6 | 6 |
| 11 | 11 |
| 10 | 10 |
| 9 | 9 |
| 9.5 | 9.5 |
| 8 | 8 |
| 9 | 9 |
| 10 | 10 |
| 10 | 10 |
| 8 | 8 |
| 8 | 8 |
| 10 | 10 |
| 10 | 10 |
| 7.5 | 7.5 |

| | |
|---|---|
| 10 | 10 |
| 10 | 10 |
| 10.5 | 10.5 |
| 11.5 | 11.5 |
| 10 | 10 |
| 9 | 9 |
| 8 | 8 |
| 10 | 10 |
| 8 | 8 |
| 10 | 10 |
| 8 | 8 |
| 7.5 | 7.5 |
| 9 | 9 |
| 8.75 | 8.75 |
| 8 | 8 |
| 10 | 10 |
| 10.5 | 10.5 |
| 8 | 8 |
| 4 | 4 |
| 10 | 10 |
| 10 | 10 |
| 10 | 10 |
| 9.5 | 9.5 |
| 8.5 | 8.5 |
| 11 | 11 |
| 11 | 11 |
| 8 | 8 |

| | |
|---|---|
| 10 | 10 |
| 8.5 | 8.5 |
| 9 | 9 |
| 6 | 6 |
| 8.5 | 8.5 |
| 8 | 8 |
| 11 | 11 |
| 10 | 10 |
| 10 | 10 |
| 9 | 9 |
| 10 | 10 |
| 10 | 10 |
| 6 | 6 |
| 10 | 10 |
| 10 | 10 |
| 11 | 11 |
| 9 | 9 |
| 10.5 | 10.5 |
| 8.5 | 8.5 |
| 10 | 10 |
| 7 | 7 |
| 10 | 10 |
| 10 | 10 |
| 12 | 12 |
| 8 | 8 |
| 10 | 10 |
| 10 | 10 |
| 7.5 | 7.5 |

| | |
|---|---|
| 7 | 7 |
| 9 | 9 |
| 11 | 11 |
| 8 | 8 |
| 7 | 7 |
| 10.5 | 10.5 |
| 10 | 10 |
| 9 | 9 |
| 10 | 10 |
| 10 | 10 |
| 8 | 8 |
| 9 | 9 |
| 11 | 11 |
| 8 | 8 |
| 10-Sep | 9.5 |
| 9.5 | 9.5 |
| 10.5 | 10.5 |
| 10 | 10 |
| 10 | 10 |
| 10 | 10 |
| 10 | 10 |
| 11 | 11 |
| 9.5 | 9.5 |
| 10 | 10 |
| 9 | 9 |
| 9.5 | 9.5 |
| 10 | 10 |
| 9 | 9 |

| | |
|---|---|
| 9.5 | 9.5 |
| 10 | 10 |
| 9.5 | 9.5 |
| 9.5 | 9.5 |
| 10 | 10 |
| 8 | 8 |
| 11 | 11 |
| 9.5 | 9.5 |
| 11 hrs | 11 |
| 8 | 8 |
| 11 | 11 |
| 11 | 11 |
| 10 | 10 |
| 6 | 6 |
| 11 | 11 |
| 8 | 8 |
| 9 | 9 |
| 10.5 | 10.5 |
| 9 | 9 |
| 10 | 10 |
| 10 | 10 |
| 10.5 | 10.5 |
| 10 | 10 |
| 8 | 8 |
| 8 hours | 8 |
| 9.5 | 9.5 |
| 10 | 10 |
| 10 | 10 |

| | |
|---|---|
| 9 | 9 |
| 8 | 8 |
| 8 | 8 |
| 9 | 9 |
| 7 | 7 |
| 10 | 10 |
| 10 | 10 |
| 9 | 9 |
| 10 | 10 |
| 10 | 10 |
| 10 | 10 |
| 9 | 9 |
| 10 | 10 |
| 8 | 8 |
| 9 | 9 |
| 10 | 10 |
| 9 | 9 |
| 10 | 10 |
| 10 | 10 |
| 9 | 9 |
| 10.5 | 10.5 |
| 11 | 11 |
| 12 | 12 |
| 9.5 | 9.5 |
| 8 | 8 |
| 9.5 | 9.5 |
| 10 | 10 |
| 8 | 8 |

| | |
|---|---|
| 11 | 11 |
| 9 | 9 |
| 9.5 | 9.5 |
| 6 | 6 |
| 9.5 | 9.5 |
| 6.5 | 6.5 |
| 9 | 9 |
| 10 | 10 |
| 9 | 9 |
| 10 | 10 |
| 8 | 8 |
| 8 | 8 |
| 8 | 8 |
| 11 | 11 |
| 8 | 8 |
| 10 | 10 |
| 10.5 | 10.5 |
| 10 | 10 |
| 11 | 11 |
| 9 | 9 |
| 9 | 9 |
| 8.5 | 8.5 |
| 8 | 8 |
| 9 | 9 |
| 11 | 11 |
| 10 | 10 |
| 8.5 | 8.5 |
| 10 | 10 |

| | |
|---|---|
| 10.5 | 10.5 |
| 9 | 9 |
| 9.5 | 9.5 |
| 9.5 | 9.5 |
| 8 | 8 |
| 9 | 9 |
| 9 | 9 |
| 10 | 10 |
| 9.5 | 9.5 |
| 7 | 7 |
| 9 | 9 |
| 9 | 9 |
| 9.5 | 9.5 |
| 10 | 10 |
| 10.5 | 10.5 |
| 10 | 10 |
| 8.5 | 8.5 |
| 10.5 | 10.5 |
| 8 | 8 |
| 9 | 9 |
| 8 | 8 |
| 9 | 9 |
| 10 | 10 |
| 10 | 10 |
| 10 | 10 |
| 10 | 10 |
| 9 | 9 |

| | |
|---|---|
| 10 | 10 |
| 6 | 6 |
| 10 | 10 |
| 7 | 7 |
| 9.5 | 9.5 |
| 9 | 9 |
| 9.5 | 9.5 |
| 8 | 8 |
| 9 | 9 |
| 12 | 12 |
| 9 | 9 |
| 11 | 11 |
| 7 | 7 |
| 8 | 8 |
| 11 | 11 |
| 8 | 8 |
| 8.5 | 8.5 |
| 9 | 9 |
| 9.5 | 9.5 |
| 10.5 | 10.5 |
| 9 | 9 |
| 8 | 8 |
| 8 | 8 |
| 10 | 10 |
| 9 | 9 |
| 10 | 10 |
| 10 | 10 |
| 8 | 8 |

| | |
|---|---|
| 9 | 9 |
| 9 | 9 |
| 10.5 | 10.5 |
| 9.5 | 9.5 |
| 9 | 9 |
| 10 | 10 |
| 9 | 9 |
| 7 | 7 |
| 9 | 9 |
| 11 | 11 |
| 8.5 | 8.5 |
| 3.5 | 3.5 |
| 8 | 8 |
| 4 | 4 |
| 11 | 11 |
| 11 | 11 |
| 6 | 6 |
| 11 | 11 |
| 8 | 8 |
| 9 | 9 |
| 9.5 | 9.5 |
| 9 | 9 |
| 7 | 7 |
| 10 | 10 |
| 9.5 | 9.5 |
| 9.5 | 9.5 |
| 10 | 10 |
| 8.5 | 8.5 |

| | |
|---|---|
| 9.5 | 9.5 |
| 10 | 10 |
| 10.5 | 10.5 |
| 12 | 12 |
| 10.5 | 10.5 |
| 6 | 6 |
| 10 | 10 |
| 8.5 | 8.5 |
| 11 | 11 |
| 9 | 9 |
| 10 | 10 |
| 9 | 9 |
| 9.5 | 9.5 |
| 7.5 | 7.5 |
| 5 | 5 |
| 10 | 10 |
| 11.5 | 11.5 |
| 9 | 9 |
| 8 | 8 |
| 8 | 8 |
| 10.5 | 10.5 |
| 8.5 | 8.5 |
| 11 | 11 |
| 8 | 8 |
| 11 | 11 |
| 8.5 | 8.5 |
| 10 | 10 |
| 10 | 10 |

| | |
|---|---|
| 11 | 11 |
| 10 | 10 |
| 8 | 8 |
| 10 | 10 |
| 10.5 | 10.5 |
| 12 | 12 |
| 9 | 9 |
| 8 | 8 |
| 9 | 9 |
| 11 | 11 |
| 7.5 | 7.5 |
| 10.5 | 10.5 |
| 9 | 9 |
| 10 | 10 |
| 10.5 | 10.5 |
| 10.5 | 10.5 |
| 10.5 | 10.5 |
| 8 | 8 |
| 9.5 | 9.5 |
| 9 | 9 |
| 7 | 7 |
| 10 | 10 |
| 9 | 9 |
| 10.5 | 10.5 |
| 9 | 9 |
| 10 | 10 |
| 11.5 | 11.5 |

| | |
|---|---|
| 9 | 9 |
| 8 | 8 |
| 9.5 | 9.5 |
| 9 | 9 |
| 10 | 10 |
| 10 | 10 |
| 11 | 11 |
| 11 | 11 |
| 10 | 10 |
| 10 | 10 |
| 8 | 8 |
| 10.5 | 10.5 |
| 10 | 10 |
| 9 | 9 |
| 10 | 10 |
| 8 | 8 |
| 11 | 11 |
| 10 | 10 |
| 10 | 10 |
| 11.5 | 11.5 |
| 10 | 10 |
| 9 | 9 |
| 10.5 | 10.5 |
| 9 | 9 |
| 9.5 | 9.5 |
| 11.5 | 11.5 |
| 11 | 11 |
| 9 | 9 |

| | |
|---|---|
| 11 | 11 |
| 10 | 10 |
| 8 | 8 |
| 10 | 10 |
| 11 | 11 |
| 10 | 10 |
| 11 | 11 |
| 10 | 10 |
| 10.5 | 10.5 |
| 9.5 | 9.5 |
| 9.5 | 9.5 |
| 10 | 10 |
| 10 | 10 |
| 8.5 | 8.5 |
| 11 | 11 |
| 8 | 8 |
| 10.5 | 10.5 |
| 9.5 | 9.5 |
| 9.5 | 9.5 |
| 11 | 11 |
| 10 | 10 |
| 8 | 8 |
| 7.5 | 7.5 |
| 9 | 9 |
| 10 | 10 |
| 9.5 | 9.5 |
| 9.5 | 9.5 |
| 11 | 11 |

| | |
|---|---|
| 10 | 10 |
| 8 | 8 |
| 7 | 7 |
| 9 | 9 |
| 10 | 10 |
| 10 | 10 |
| 11 | 11 |
| 9.5 | 9.5 |
| 10 | 10 |
| 11 | 11 |
| 7.5 | 7.5 |
| 10 | 10 |
| 10 | 10 |
| 8 | 8 |
| 10 | 10 |
| 10 | 10 |
| 10 | 10 |
| 11 | 11 |
| 11 | 11 |
| 8 | 8 |
| 9 | 9 |
| 8.5 | 8.5 |
| 7 | 7 |
| 9 | 9 |
| 11 | 11 |
| 9 | 9 |
| 8 | 8 |
| 10 | 10 |

| | |
|---|---|
| 9.5 | 9.5 |
| 8 | 8 |
| 8 | 8 |
| 9 | 9 |
| 11 | 11 |
| 5 | 5 |
| 10 | 10 |
| 9 | 9 |
| 10 | 10 |
| 10 | 10 |
| 10 | 10 |
| 8 | 8 |
| 9 | 9 |
| 10 | 10 |
| 9.5 | 9.5 |
| 10 | 10 |
| 11 | 11 |
| 9.5 | 9.5 |
| 11 | 11 |
| 9 | 9 |
| 9 | 9 |
| 8 | 8 |
| 8 | 8 |
| 12 | 12 |
| 9 | 9 |
| 10 | 10 |
| 9.5 | 9.5 |
| 10 | 10 |

| | |
|---|---|
| 8 | 8 |
| 9 | 9 |
| 8 | 8 |
| 10 | 10 |
| 9 | 9 |
| 7 | 7 |
| 9.5 | 9.5 |
| 9 | 9 |
| 8 | 8 |
| 10 | 10 |
| 8 | 8 |
| 8.5 | 8.5 |
| 9 | 9 |
| 11 | 11 |
| 8.5 | 8.5 |
| 11 | 11 |
| 10 | 10 |
| 10 | 10 |
| 10 | 10 |
| 8 | 8 |
| 8.5 | 8.5 |
| 8 | 8 |
| 10.5 | 10.5 |
| 4.5 | 4.5 |
| 10 | 10 |
| 8 | 8 |
| 10 | 10 |
| 9.5 | 9.5 |

| | |
|---|---|
| 10.5 | 10.5 |
| 10 | 10 |
| 8.5 | 8.5 |
| 10 | 10 |
| 11 | 11 |
| 9.5 | 9.5 |
| 9 | 9 |
| 9.5 | 9.5 |
| 8 | 8 |
| 9 | 9 |
| 12 | 12 |
| 10.5 | 10.5 |
| 10.5 | 10.5 |
| 6 | 6 |
| 8 | 8 |
| 10.5 | 10.5 |
| 8 | 8 |
| 9 | 9 |
| 5 | 5 |
| 10 | 10 |
| 9 | 9 |
| 10 | 10 |
| 9.5 | 9.5 |
| 8.5 | 8.5 |
| 8.5 | 8.5 |
| 6 | 6 |
| 10 | 10 |
| 8 | 8 |

| | |
|---|---|
| 10 | 10 |
| 8 | 8 |
| 10 | 10 |
| 8.5 | 8.5 |
| 10.5 | 10.5 |
| 10.5 | 10.5 |
| 10 | 10 |
| 5.5 | 5.5 |
| 8 | 8 |
| 11 | 11 |
| 10 | 10 |
| 9.75 | 9.75 |
| 10 | 10 |
| 10 | 10 |
| 11 | 11 |
| 8 | 8 |
| 11 | 11 |
| 8.5 | 8.5 |
| 9 | 9 |
| 10.5 | 10.5 |
| 9 | 9 |
| 4 | 4 |
| 10 | 10 |
| 8 | 8 |
| 10.5 | 10.5 |
| 12 | 12 |
| 8 | 8 |

| | |
|---|---|
| 10 | 10 |
| 8 | 8 |
| 10 | 10 |
| 8 | 8 |
| 10 | 10 |
| 9.5 | 9.5 |
| 6 | 6 |
| 7 | 7 |
| 7 | 7 |
| 11.5 | 11.5 |
| 8 | 8 |
| 8 | 8 |
| 11 | 11 |
| 10 | 10 |
| 9 | 9 |
| 12 | 12 |
| 11 | 11 |
| 9 | 9 |
| 10 | 10 |
| 9 | 9 |
| 8 | 8 |
| 8 | 8 |
| 7 | 7 |
| 10 | 10 |
| 9 | 9 |
| 8.5 | 8.5 |
| 8 | 8 |
| 9 | 9 |

| | |
|---|---|
| 6 | 6 |
| 8 | 8 |
| 8 | 8 |
| 8 | 8 |
| 10 | 10 |
| 8 | 8 |
| 11 | 11 |
| 10 | 10 |
| 6 | 6 |
| 10 | 10 |
| 11 | 11 |
| 8 | 8 |
| 8 | 8 |
| 8 | 8 |
| 8 | 8 |
| 10 | 10 |
| 11 | 11 |
| 8 | 8 |
| 7.5 | 7.5 |
| 11 | 11 |
| 8.5 | 8.5 |
| 8 | 8 |
| 8 | 8 |
| 9 | 9 |
| 10 | 10 |
| 9 | 9 |
| 10 | 10 |

| | |
|---|---|
| 10 | 10 |
| 10 | 10 |
| 5 | 5 |
| 9 | 9 |
| 10 | 10 |
| 9.5 | 9.5 |
| 10 | 10 |
| 10 | 10 |
| 7.5 | 7.5 |
| 9 | 9 |
| 10 | 10 |
| 10 | 10 |
| 10.5 | 10.5 |
| 10 | 10 |
| 10 | 10 |
| 9 | 9 |
| 9 | 9 |
| 8 | 8 |
| 8 | 8 |
| 10.5 | 10.5 |
| 10 | 10 |
| 9 | 9 |
| 11 | 11 |
| 9.5 | 9.5 |
| 10 | 10 |
| 10 | 10 |
| 10 | 10 |

| | |
|---|---|
| 9.5 | 9.5 |
| 10 | 10 |
| 12 | 12 |
| 8 | 8 |
| 9 | 9 |
| 10 | 10 |
| 9 | 9 |
| 6 | 6 |
| 12 | 12 |
| 7.5 | 7.5 |
| 9.5 | 9.5 |
| 10 | 10 |
| 8 | 8 |
| 10 | 10 |
| 10 | 10 |
| 10 | 10 |
| 9 | 9 |
| 10 | 10 |
| 10 | 10 |
| 9 | 9 |
| 9.5 | 9.5 |
| 10 | 10 |
| 10.5 | 10.5 |
| 9.5 | 9.5 |
| 9.5 | 9.5 |
| 8 | 8 |
| 9 | 9 |
| 12 | 12 |

| | |
|---|---|
| 9 | 9 |
| 9.5 | 9.5 |
| 8 | 8 |
| 8 | 8 |
| 8 | 8 |
| 9 | 9 |
| 11 | 11 |
| 9 | 9 |
| 8 | 8 |
| 9.5 | 9.5 |
| 11 | 11 |
| 9 | 9 |
| 8 | 8 |
| 9 | 9 |
| 9 | 9 |
| 10 | 10 |
| 10 | 10 |
| 10 | 10 |
| 8.5 | 8.5 |
| 10 | 10 |
| 8.5 | 8.5 |
| 9 | 9 |
| 8 | 8 |
| 10 | 10 |
| 10 | 10 |
| 9.5 | 9.5 |
| 7 | 7 |
| 8.5 | 8.5 |

| | |
|---|---|
| 8 | 8 |
| 6 | 6 |
| 8 | 8 |
| 9 | 9 |
| 10 | 10 |
| 10.5 | 10.5 |
| 10 | 10 |
| 8 | 8 |
| 10 | 10 |
| 9 | 9 |
| 10 | 10 |
| 11 | 11 |
| 7 | 7 |
| 9 | 9 |
| 9 | 9 |
| 11 | 11 |
| 8 | 8 |
| 8 | 8 |
| 9 | 9 |
| 12.5 | 12.5 |
| 10 | 10 |
| 9 | 9 |
| 7 | 7 |
| 9.5 | 9.5 |
| 9 | 9 |
| 7.5 | 7.5 |
| 10 | 10 |
| 9.5 | 9.5 |

| | |
|---|---|
| 9.5 | 9.5 |
| 6 | 6 |
| 9 | 9 |
| 10.5 | 10.5 |
| 10 | 10 |
| 10 | 10 |
| 10 | 10 |
| 10 | 10 |
| 11 | 11 |
| 11 | 11 |
| 10 | 10 |
| 6.5 | 6.5 |
| 10 | 10 |
| 8.5 | 8.5 |
| 8 | 8 |
| 8 | 8 |
| 8.5 | 8.5 |
| 12 | 12 |
| 8 | 8 |
| 9.5 | 9.5 |
| 10 | 10 |
| 10 | 10 |
| 9 | 9 |
| 10 | 10 |
| 9.5 | 9.5 |
| 10.5 | 10.5 |
| 10 | 10 |
| 10 | 10 |

| | |
|---|---|
| 9.5 | 9.5 |
| 10 | 10 |
| 8 | 8 |
| 11 | 11 |
| 11 | 11 |
| 9 | 9 |
| 9 | 9 |
| 9 | 9 |
| 7 | 7 |
| 11 | 11 |
| 10 | 10 |
| 12 | 12 |
| 8 | 8 |
| 9 | 9 |
| 10 | 10 |
| 10 | 10 |
| 11 | 11 |
| 9 | 9 |
| 9 | 9 |
| 10 | 10 |
| 6.5 | 6.5 |
| 9 | 9 |
| 9.5 | 9.5 |
| 10 | 10 |
| 7.5 | 7.5 |
| 10 | 10 |

| | |
|---|---|
| 9 | 9 |
| 9.5 | 9.5 |
| 10 | 10 |
| 9.5 | 9.5 |
| 10.5 | 10.5 |
| 11 | 11 |
| 7 | 7 |
| 10 | 10 |
| 10 | 10 |
| 10 | 10 |
| 8 | 8 |
| 9 | 9 |
| 9 | 9 |
| 12 | 12 |
| 9.5 | 9.5 |
| 9 | 9 |
| 10.5 | 10.5 |
| 10 | 10 |
| 11 | 11 |
| 8 | 8 |
| 10.5 | 10.5 |
| 7 | 7 |
| 10 | 10 |
| 10 | 10 |
| 7 | 7 |
| 11 | 11 |
| 8 | 8 |

| | |
|---|---|
| 10 | 10 |
| 10.5 | 10.5 |
| 9 | 9 |
| 10 | 10 |
| 8 | 8 |
| 9 | 9 |
| 10 | 10 |
| 10 | 10 |
| 8 | 8 |
| 8 | 8 |
| 10 | 10 |
| 10 | 10 |
| 8 | 8 |
| 12 | 12 |
| 8 | 8 |
| 8.5 | 8.5 |
| 10 | 10 |
| 10 | 10 |
| 9 | 9 |
| 10 | 10 |
| 10 | 10 |
| 10 | 10 |
| 8 | 8 |
| 12 | 12 |
| 7 | 7 |
| 9 | 9 |
| 10 | 10 |
| 9 | 9 |

| | |
|---|---|
| 9 | 9 |
| 9 | 9 |
| 10 | 10 |
| 12 | 12 |
| 8 | 8 |
| 10 | 10 |
| 9 | 9 |
| 9.5 | 9.5 |
| 8 | 8 |
| 8.5 | 8.5 |
| 10.5 | 10.5 |
| 8 | 8 |
| 10.5 | 10.5 |
| 10 | 10 |
| 10 | 10 |
| 10 | 10 |
| 8.5 | 8.5 |
| 9 | 9 |
| 9 | 9 |
| 10 | 10 |
| 10 | 10 |
| 10 | 10 |
| 10 | 10 |
| 9.5 | 9.5 |
| 7 | 7 |
| 12 | 12 |
| 10 | 10 |
| 12 | 12 |

| | |
|---|---|
| 9 | 9 |
| 9 | 9 |
| 8 | 8 |
| 9 | 9 |
| 10 | 10 |
| 10 | 10 |
| 10 | 10 |
| 8.5 | 8.5 |
| 8 | 8 |
| 9.5 | 9.5 |
| 8 | 8 |
| 8 | 8 |
| 6 | 6 |
| 8 | 8 |
| 11.5 | 11.5 |
| 11 | 11 |
| 9 | 9 |
| 10 | 10 |
| 8 | 8 |
| 8 | 8 |
| 10 | 10 |
| 9 | 9 |
| 9.5 | 9.5 |
| 10 | 10 |
| 9 | 9 |
| 9 | 9 |

| | |
|---|---|
| 10.5 | 10.5 |
| 10 | 10 |
| 10 | 10 |
| 11 | 11 |
| 9 | 9 |
| 9 | 9 |
| 10.5 | 10.5 |
| 9.5 | 9.5 |
| 8 | 8 |
| 10.5 | 10.5 |
| 9 | 9 |
| 8.5 | 8.5 |
| 9 | 9 |
| 7.5 | 7.5 |
| 9 | 9 |
| 10 | 10 |
| 11.5 | 11.5 |
| 9 | 9 |
| 10.5 | 10.5 |
| 9 | 9 |
| 8 | 8 |
| 9 | 9 |
| 8.5 | 8.5 |
| 10 | 10 |
| 8.5 | 8.5 |
| 9 | 9 |
| 8.5 | 8.5 |

| | |
|---|---|
| 8 | 8 |
| 8.5 | 8.5 |
| 11 | 11 |
| 8.5 | 8.5 |
| 7.5 | 7.5 |
| 10 | 10 |
| 7 | 7 |
| 9 | 9 |
| 10 | 10 |
| 12 | 12 |
| 9 | 9 |
| 9 | 9 |
| 8.5 | 8.5 |
| 9 | 9 |
| 10 | 10 |
| 13 | 13 |
| 11 | 11 |
| 8 | 8 |
| 10 | 10 |
| 10 | 10 |
| 11 | 11 |
| 10 | 10 |
| 5 | 5 |
| 9 | 9 |
| 6.5 | 6.5 |
| 11 | 11 |
| 11 | 11 |
| 10 | 10 |

| | |
|---|---|
| 8 | 8 |
| 8.5 | 8.5 |
| 8 | 8 |
| 10 | 10 |
| 9.25 | 9.25 |
| 10 | 10 |
| 10 | 10 |
| 9.5 | 9.5 |
| 8 | 8 |
| 10 | 10 |
| 9 | 9 |
| 9 | 9 |
| 10 | 10 |
| 9 | 9 |
| 7 | 7 |
| 10 | 10 |
| 8.5 | 8.5 |
| 8 | 8 |
| 9 | 9 |
| 9 | 9 |
| 10 | 10 |
| 5 | 5 |
| 10.5 | 10.5 |
| 10 | 10 |
| 8.5 | 8.5 |
| 9 | 9 |
| 9.5 | 9.5 |
| 7.5 | 7.5 |

| | |
|---|---|
| 9 | 9 |
| 8 | 8 |
| 8.5 | 8.5 |
| 8 | 8 |
| 8 | 8 |
| 9 | 9 |
| 8 | 8 |
| 8 | 8 |
| 10 | 10 |
| 8 | 8 |
| 9.5 | 9.5 |
| 7 | 7 |
| 10 | 10 |
| 11 | 11 |
| 9.5 | 9.5 |
| 10 | 10 |
| 10 | 10 |
| 10 | 10 |
| 7 | 7 |
| 8 | 8 |
| 8 | 8 |
| 10 | 10 |
| 6 | 6 |
| 9 | 9 |
| 9 | 9 |
| 8 | 8 |
| 8.5 | 8.5 |
| 9 | 9 |

| | |
|---|---|
| 9 | 9 |
| 8 | 8 |
| 10 | 10 |
| 7.5 | 7.5 |
| 7 | 7 |
| 10 | 10 |
| 9 | 9 |
| 10 | 10 |
| 11 | 11 |
| 11 | 11 |
| 9.5 | 9.5 |
| 8 | 8 |
| 8 | 8 |
| 7 | 7 |
| 10 | 10 |
| 6.5 | 6.5 |
| 8 | 8 |
| 9.5 | 9.5 |
| 9 | 9 |
| 10 | 10 |
| 8 | 8 |
| 9 | 9 |
| 12 | 12 |
| 9 | 9 |
| 6 | 6 |
| 10 | 10 |
| 8 | 8 |
| 6 | 6 |

| | |
|---|---|
| 9 | 9 |
| 8 | 8 |
| 10 | 10 |
| 7 | 7 |
| 11 | 11 |
| 9 | 9 |
| 7 | 7 |
| 10 | 10 |
| 9 | 9 |
| 6 | 6 |
| 8 | 8 |
| 10 | 10 |
| 10 | 10 |
| 9 | 9 |
| 5.5 | 5.5 |
| 11 | 11 |
| 10 | 10 |
| 10 | 10 |
| 11 | 11 |
| 9 | 9 |
| 7 | 7 |
| 11.5 | 11.5 |
| 8 | 8 |
| 9 | 9 |
| 9 | 9 |
| 8 | 8 |
| 8 | 8 |
| 8 | 8 |

| | |
|---|---|
| 9.5 | 9.5 |
| 8.5 | 8.5 |
| 8 | 8 |
| 9 | 9 |
| 9.5 | 9.5 |
| 7 | 7 |
| 9.5 | 9.5 |
| 9 | 9 |
| 9.5 | 9.5 |
| 10 | 10 |
| 12 | 12 |
| 10 | 10 |
| 8 | 8 |
| 9 | 9 |
| 10 | 10 |
| 9 | 9 |
| 9 | 9 |
| 9 | 9 |
| 9 | 9 |
| 6.5 | 6.5 |
| 10.5 | 10.5 |
| 7 | 7 |
| 10 | 10 |
| 9 | 9 |
| 11 | 11 |
| 11 | 11 |
| 9 | 9 |

| | |
|---|---|
| 10 | 10 |
| 8 | 8 |
| 10 | 10 |
| 8 | 8 |
| 10 | 10 |
| 8.5 | 8.5 |
| 8.5 | 8.5 |
| 9 | 9 |
| 9.5 | 9.5 |
| 9 | 9 |
| 8 | 8 |
| 8.5 | 8.5 |
| 9 | 9 |
| 10 | 10 |
| 9 | 9 |
| 11 | 11 |
| 11 | 11 |
| 6 | 6 |
| 9 | 9 |
| 11 | 11 |
| 10 | 10 |
| 10 | 10 |
| 6 | 6 |
| 9.5 | 9.5 |
| 8 | 8 |
| 10 | 10 |
| 8 | 8 |
| 9 | 9 |

| | |
|---|---|
| 9 | 9 |
| 10 | 10 |
| 9 | 9 |
| 11 | 11 |
| 9.5 | 9.5 |
| 8.5 | 8.5 |
| 10 | 10 |
| 8 | 8 |
| 10 | 10 |
| 8 | 8 |
| 8.5 | 8.5 |
| 10 | 10 |
| 10 | 10 |
| 10 | 10 |
| 7 | 7 |
| 9 | 9 |
| 8 | 8 |
| 10 | 10 |
| 8.5 | 8.5 |
| 6.5 | 6.5 |
| 3 | 3 |
| 6.5 | 6.5 |
| 10 | 10 |
| 9.5 | 9.5 |
| 7.5 | 7.5 |
| 8 | 8 |
| 8.5 | 8.5 |
| 10 | 10 |

| | |
|---|---|
| 10 | 10 |
| 10 | 10 |
| 10 | 10 |
| 11 | 11 |
| 9 | 9 |
| 9.5 | 9.5 |
| 8 | 8 |
| 11 | 11 |
| 7.5 | 7.5 |
| 9.5 | 9.5 |
| 9.5 | 9.5 |
| 10.5 | 10.5 |
| 8 | 8 |
| 9 | 9 |
| 9 | 9 |
| 7 | 7 |
| 8 | 8 |
| 10.5 | 10.5 |
| 9 | 9 |
| 11 | 11 |
| 10.5 | 10.5 |
| 10 | 10 |
| 7.5 | 7.5 |
| 9 | 9 |
| 9 | 9 |
| 9 | 9 |
| 10 | 10 |
| 8.5 | 8.5 |

| | |
|---|---|
| 7 | 7 |
| 11 | 11 |
| 8 | 8 |
| 9 | 9 |
| 11 | 11 |
| 9 | 9 |
| 10 | 10 |
| 9.5 | 9.5 |
| 7 | 7 |
| 7.5 | 7.5 |
| 10 | 10 |
| 8 | 8 |
| 8 | 8 |
| 8.5 | 8.5 |
| 10 | 10 |
| 8.5 | 8.5 |
| 9 | 9 |
| 10 | 10 |
| 11 | 11 |
| 9.5 | 9.5 |
| 8 | 8 |
| 11 | 11 |
| 10 | 10 |
| 9 | 9 |
| 9 | 9 |

| | |
|---|---|
| 8 | 8 |
| 9 | 9 |
| 10 | 10 |
| 9 | 9 |
| 10 | 10 |
| 7.5 | 7.5 |
| 11 | 11 |
| 8 | 8 |
| 10 | 10 |
| 6 | 6 |
| 10 | 10 |
| 9.5 | 9.5 |
| 9 | 9 |
| 10 | 10 |
| 7.5 | 7.5 |
| 10 | 10 |
| 8 | 8 |
| 7 | 7 |
| 8.5 | 8.5 |
| 9 | 9 |
| 11 | 11 |
| 10 | 10 |
| 10 | 10 |
| 10 | 10 |
| 9 | 9 |
| 9 | 9 |
| 10 | 10 |

| | |
|---|---|
| 8.5 | 8.5 |
| 9 | 9 |
| 9 | 9 |
| 9.5 | 9.5 |
| 8 | 8 |
| 9 | 9 |
| 10 | 10 |
| 5 | 5 |
| 11.5 | 11.5 |
| 10 | 10 |
| 11 | 11 |
| 9 | 9 |
| 8 | 8 |
| 12 | 12 |
| 9 | 9 |
| 9.5 | 9.5 |
| 8 | 8 |
| 11 | 11 |
| 10 | 10 |
| 9 | 9 |
| 8.5 | 8.5 |
| 11 | 11 |
| 9 | 9 |
| 9 | 9 |
